# A Cell-Based Papain-like Protease (PLpro) Activity Assay for Rapid Detection of Active SARS-CoV-2 Infections and Antivirals

**DOI:** 10.1101/2024.05.21.24307588

**Authors:** Anahi G. Jimenez-Campos, Lucas I. Maestas, Nileena Velappan, Brian Beck, Chunyan Ye, Keith Wernsing, Yaniksa Mata-Solis, William J. Bruno, Silas C. Bussmann, Steven Bradfute, Justin T. Baca, Frauke H. Rininsland

## Abstract

Severe acute respiratory syndrome coronavirus 2 (SARS-CoV-2) and its variants are a continuous threat to human life and prosperity. An urgent need remains for simple and fast tests that reliably detect active infections with SARS-CoV-2 and its variants in asymptomatic and presymptomatic carriers. Here we introduce a simple and rapid activity-based diagnostic (ABDx) test that identifies SARS-CoV-2 infections by measuring the activity of a viral enzyme, Papain-Like protease (PLpro). The test system consists of a peptide that fluoresces when cleaved by SARS PLpro that is active in crude, unprocessed lysates from human tongue scrapes and saliva. Test results are obtained in 30 minutes or less using widely available fluorescence plate readers, or a battery-operated portable instrument for testing at point of care (PoC) sites and in low-resource and hard to reach populations. Proof-of-concept was obtained in a clinical study on clinical specimens collected from patients with COVID-19 like symptoms who tested positive (n=10) or negative (n=10) with diagnostic LIAT RT-PCR using nasal mid turbinate swabs. When saliva from these patients was tested with in-house endpoint RT-PCR only 3 specimens were negative and 17 were positive. PLpro activity correlated in 17 of these cases (3 out of 3 negatives and 14 out of 16 positives, with one invalid specimen. Despite the small number of samples, the agreement was significant (p value = 0.01). In one presumptive false positive patient viral RNA was detected in saliva 5 days later by endpoint RT-PCR. Two false negatives were detected, one from a sample with a late Ct value of 35 in diagnostic RT-PCR, indicating that an active infection was no longer present. The PLpro activity assay is easily scalable and expected to detect all viable SARS-CoV-2 variants and recombinants, making it attractive as a screening and surveillance tool. Additionally, we show feasibility of the platform as a new homogeneous phenotypic assay for rapid screening of SARS-CoV-2 antiviral drugs and neutralizing antibodies.

## Introduction

Since the emergence of the severe acute respiratory syndrome coronavirus 2 (SARS-CoV-2) in 2019, over 2,200 different diagnostic tests have been developed and introduced to the market targeting either SARS-CoV-2 nucleic acids or antigens.^1^ The SARS-CoV-2 Papain-Like protease (PLpro) and the main protease (MPro, also known as 3CLPro) present a third class of detection target. PLpro and MPro cleave the viral polyprotein at conserved sites to separate the non-structural proteins nsp 1-3 and nsp 4-16, respectively.^2,3^ The proteolytic activities can be measured using synthetic peptides that are specifically recognized by the protease of interest and fluoresce when cleaved. This kind of assay belongs to a category known as activity-based diagnostics (ABDx) and are gaining recognition as important methodologies for detecting pathogen infections in clinical specimens.^4,5^ Measuring SARS-CoV-2 protease activities has several advantages.

### ABDx assays detect only active infections

SARS-CoV-2 proteases are expressed only in infected cells that are actively assembling new virions; therefore, measuring viral protease activity is likely to accurately identify only those individuals able to transmit infectious virus. In contrast, RT-PCR cannot differentiate between replication competent virus and residual, non-infectious RNA that in some cases has been detected 28 days or even months after symptoms have cleared.^6,7^ Currently, the presence of infectious virus is assessed in cell culture by determining the cytopathic effect (CPE), detecting viral fragments by RT-PCR or immunostaining, using plaque assays, focus-forming assays, or by determining the 50% tissue culture infectious dose (TCID_50_).^8,9^ These methods cannot be used for routine specimen testing since they are time-and labor intensive and restricted to biosafety level 3 conditions. Negative sense SARS-CoV-2 RNA, which is produced only by replicating virus, can also be used as an indicator of infectiousness, but this type of testing is not inexpensive or rapid, and therefore not suitable for population screening.^10^ Due to these limitations, at the beginning of the pandemic a patient’s ability to transmit virus was predicted by the arbitrary test-specific cycle threshold (Ct) value determined in RT-PCR, but this approach has since proved unreliable. ^11–14^

### Protease activities are detectable early in an infection

After SARS-CoV-2 has entered a cell, intracellular virions are produced within 10 hours but typically not released from the cell until 72 hours postinfection.^15,16^ Since the SARS-CoV-2 proteases PLpro and MPro are required for the production of intracellular virions, it is expected that their activities are detectable in infected cells several days before RNA is collectable from extracellular virions that are shed into specimens. As a result, RT-PCR tests typically perform best 2 days after symptom onset when viral loads are highest, missing pre-symptomatic and asymptomatic transmission.^17^ One study suggests a sensitivity approaching 0 % corresponding to a false negative rate of nearly 100 % at the early stages of infection, including periods when at least some people are already infectious.^18^ Early detection of infection by measuring protease activities would facilitate the rapid isolation of infected people and early treatment of the vulnerable population.

### Detection of protease activities is likely “variant proof”

SARS-CoV-2 proteases do not tolerate mutations in the catalytic domains that would interfere with their cleavage activities and thereby cause a false negative test result with an ABDx test. A recent analysis of 70,000 PLpro sequences identified mutations in only 5%, all in regions outside of the catalytically active domains.^19^ It is therefore highly likely that PLpro activity is measurable in any of the continuously emerging SARS-CoV-2 variants that may be missed by RT-PCR when primers or probes fail to bind to mutated sequences. One study reported that 4-5 individual variants may coexist in one person in which 79% of primer binding sequences in at least 1 gene were mutated, with the result that 10-15% of variants were not detectable.^20–22^

### Cell-based protease activity assays for screening of SARS-CoV-2 antivirals

Although both SARS-CoV-2 MPro and PLpro are attractive drug targets, so far only one targeted therapy—the MPro inhibitor nirmatrelvir (a component of Paxlovid)—has received approval for use in the United States.^23,24^ Despite the considerable efforts that have been invested in the development of PLpro inhibitors, promising lead candidates have not progressed to animal models. One possible reason is that many studies have used only the minimal proteolytic PLpro domain to screen for compound inhibition in biochemical screens, neglecting the effects of other SARS-CoV-2 domains involved in regulating the activity of PLpro.^25^ Although these limitations can be overcome using phenotypic assays, these tests require several steps and 5-7 days to measure the effect of drugs on cell proliferation or health.

Further, measuring SARS-CoV-2 protease activity as a marker of infection allows to rapidly determine the efficacy of therapeutic or vaccine-derived antibodies. Neutralization assays, such as plaque reduction neutralization test (PRNT), fluorescent antibody virus neutralization (FAVN) assays and immunological assays, play a pivotal role in evaluating the efficacy of vaccines, selecting convalescent plasma for clinical trials and therapeutic use, and defining immune evasion.^26^ However, those assays require several steps and days to complete.^26^ In contrast, cell-based protease activity assays are simple and can be automated and completed in one working day with minimal user steps.

We report here on an ABDx platform optimized for detecting the activity of SARS-CoV-2 Papain-like protease (PLpro) in crude, active lysates from infected cells. We show proof of concept as a Point of Care (PoC) diagnostic test by demonstrating the ability to detect SARS-CoV-2 PLpro activity in unprocessed lysates from patient saliva or tongue scrape specimens in a clinical feasibility study. A second study showed high percent agreement with endpoint RT-PCR in saliva specimens. Additional data highlight the potential of the platform as a faster and safer alternative to current phenotypic assays that could accelerate the discovery of anti-viral drugs and neutralizing antibodies.

## MATERIALS AND METHODS

### Assay Principle

We used florescence “turn on” assays for SARS-CoV-2 PLpro and Mpro in which lysates are incubated with a peptide labeled with a fluorophore at the terminal amino acid. In the intact peptide, the fluorescence is quenched by the electrons in the amide bonds and restored when the protease cleaves the bond between the terminal amino acid of the peptide and the fluorophore (Figure 1).

**Figure 1.**
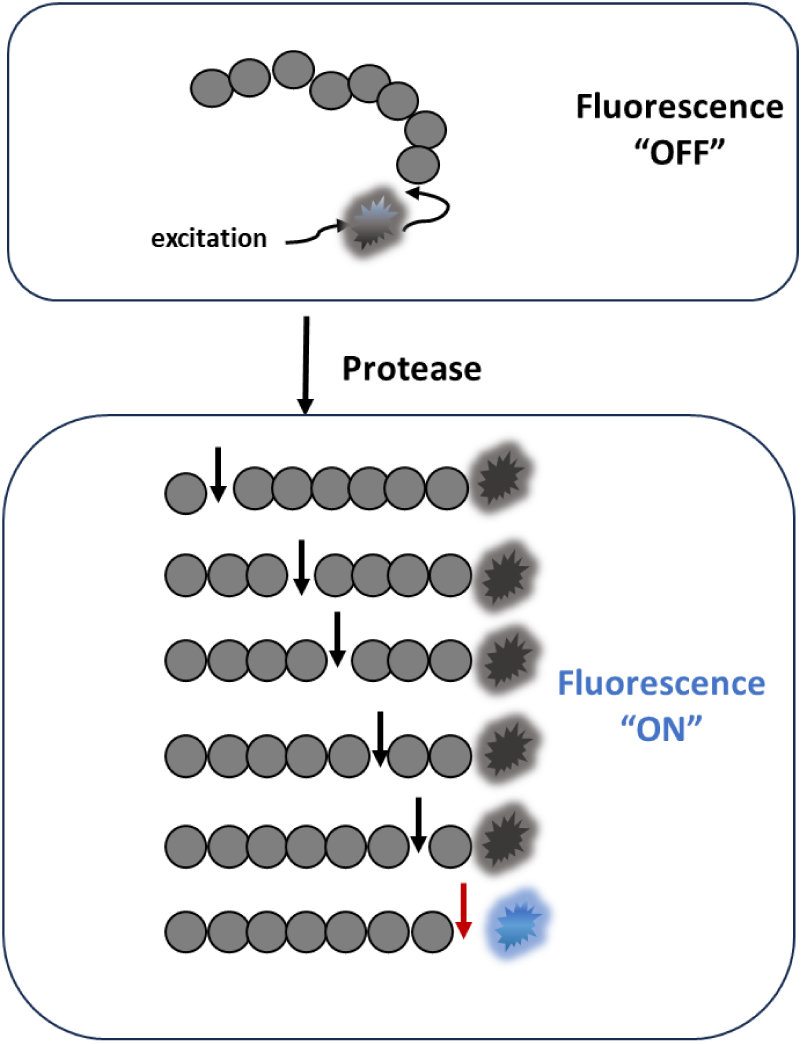
Schematic of the SARS-CoV-2 protease fluorescence turn on assay. The assay uses peptides composed of amino acids (grey circles) that are cleaved by the protease of interest and a fluorophore (starburst) conjugated to the terminal amino acid. In the intact peptide, the fluorescence is quenched by the electron withdrawing property of the amide bond. Fluorescence is restored (blue starburst) only when cleavage occurs at the bond between the terminal amino acid and the fluorophore (red arrow). Cleavage between amino acids (black arrows) by other proteases that may be present in a crude lysate does not result in notable fluorescence (grey starburst). Peptides used in this study were labeled at the carboxy terminus with rhodamine 110 (R110) or amino 7-amino-4-methylcoumarin (AMC).

#### Peptides

PLpro peptide substrates consisted of various stretches of amino acids from human or mouse ISG15 and the known terminal PLpro cleavage sequence LRGG. Peptides P4-AMC and P16-AMC were 4 and 16 amino acids in length and labeled with 7-amino-4-methylcoumarin (AMC). P4-AMC was purchased from BioTechne, (Minneapolis, MN) and P16-AMC synthesized by LifeTein (Somerset, NJ). PLpro peptides containing unnatural amino acids in the cleavage sequence (denoted as “Pun”) were used in cell-based assays. The peptides Pun16 and Pun74 were 16 and 74 amino acids in length and synthesized and labeled with AMC or rhodamine 110 (R110) at the carboxy terminus by LifeTein and AmbioPharm (Beech Island, SC). ISG15-R110 was purchased from SouthBay BioScience (San Francisco, CA) or from UBPBio (Amsterdam, The Netherlands), ISG15-AMC was from R&D Systems and ubiquitin-AMC from UBPBio.

The MPro peptide with the sequence LGSAVLQ-rhodamine110-dP was purchased from R&D Systems (Minneapolis, MN). This peptide is cleaved after glutamine (Q) by MPro from coronaviruses but rarely by other human proteases.^27^

Internal control peptides were used to demonstrate that lysates were of sufficient quality and quantity for the assay to perform. The peptide with the sequence FITC-YVADAP-DNP (Bachem, Vista, CA) is cleaved by the Angiotensin Converting Enzyme 2 (ACE2) and the peptide with the sequence DEVD by caspases. DEVD peptides were labeled with AMC or R110 and purchased from R&D Systems (Minneapolis, MN).

#### Enzymes

Recombinant papain-like proteases (PLPs) from SARS-1, SARS-CoV-2, NL63 and caspase 3 were purchased from BPS BioScience (San Diego, CA). Recombinant USP18 was obtained from Boston Biochemicals and AdenoL3 protease from MyBiosource (San Diego, CA, #MBS1182235_61). Recombinant PLPs from MERS, OC43, 229E and HKU1 were produced by Genscript (Piscataway, NJ) and contained amino acids spanning the active site and UBL-1 sites. MERS (Genbank accession number AFS88944) contained amino acids 1482-1802, OC43 (Genbank accession number AMK59674) amino acids 1562-1870, 229E (Genbank accession number APT69896) amino acids 1590-1904, and HKU1 (Genbank accession number ARB07606) amino acids 1642-1957. Genes were synthesized and codon optimized, cloned into the EcoR1/HindIII cloning sites of the pcDNA3.4 vector, transfected into Chinese Hamster Ovary cells (CHO-S), and purified via HIS tag according to protocols established at Genscript. The quality of the proteins was confirmed by SDS-PAGE, Western blot and analytical SEC-HPLC.

### Biochemical enzyme activity assays

#### Papain-like proteases

Biochemical assays using recombinant proteases were performed to optimize SARS-CoV-2 PLpro assay conditions, characterize, and optimize PLpro substrates, and to test for cross reactive cleavage by other papain-like proteases. Assays were performed in 20µl assay buffer (100mM HEPES, 5mM DTT, pH 7.4) using substrate concentrations ranging from 2µM to 20µM. The final concentrations of recombinant PLPs from SARS-CoV and SARS-CoV-2 were 100nM, NL63 428nM, MERS 1178nM, OC43 1760nM, 229E 1200nM and HKU1 1200nM. The fluorescence from cleaved AMC or R110 was measured using excitation and emission wavelengths of ʎ_ex_=365 nm and ʎ_em_=465 nm for AMC and ʎ_ex_=485 nm and ʎ_em_=528 nm for R110-labeled substrates in a fluorescence plate reader (Synergy H1, Agilent Technologies, Santa Clara, California) in white 384-well plates (Corning Inc., Corning, NY).

#### MPro

MPro assays were performed using the same assay buffer as for PLpro, with 10µM peptide and between 5 and 500 nM enzyme.

#### Caspase-3

Internal control assays were performed using the assay buffer as above, 1.5nM caspase 3, and 10µM peptide (DEVD_2_-R110). In some assays, PLpro and caspase peptides labeled with different fluorophores were combined and monitored in one reaction using dual wavelength monitoring for AMC and R110.

#### USP18

USP18 (1000 nM) was reacted with human ISG15-R110 (500nM) in assay buffer containing 50 mM HEPES, pH 7.5, 10 mM DTT, 0.01% Tween.

#### Adenovirus protease L3

Enzyme (250nM) was reacted with 66µM peptide P4 or Pun4 in 25mM TRIS-HCL, pH 7.4, 2mM DTT, 0.3% octyglucoside.

#### Sample Quantification

(“Quanti Dye”). The nucleic acid content in lysates was measured to estimate the amount of biological material collected in tongue scrape and saliva specimens. The nucleic acid intercalating dye SYBR® Safe Stain (Edvotek, Washington, DC) was used at a 2.5x dilution in 100mM HEPES, pH 7.4. The linear region of detection was determined by adding QuantiDye to triplicate wells of crude lysate that was diluted in steps of 1:2 in lysis buffer. The linear detection range was between 74,000 RFU to > 1.5 million RFU (Figure 1, Supplemental Information).

### HCoV NL63, OC43, 229E infection and activity assays

All experiments with HCoV propagation were performed in biosafety level 2 containment laboratories at Los Alamos National Laboratory with approved protocols. MRC-5 cells (ATCC, CCL-171), HCT8 cells (ATCC, CCL-244), and LLC-MK2 cells (ATCC, CCL-7) were grown to 90% confluency (approx. 1× 10^5^ cells) in 12-well plates in Dulbecco’s minimal essential medium (DMEM) supplemented with 10% fetal calf serum (FCS) and 1% penicillin/streptomycin. MRC-5 cells were infected with 2 MOI of 229E (ATCC, VR-740) for 24 hours, HCT-8 with 2 MOI of OC43 (ATCC, VR-1558) for 5 days, and LLC-MK2 with 1 MOI of NL63 (BEI NR-470) for 24 hours in viral growth medium (MEM + 2.5% heat inactivated fetal calf serum) at 37°C in a humidified incubator.

#### HCoV OC43, 229E and NL63 protease activity assays

After cells were incubated with virus, the media was removed from the wells and cells washed three times with 100mM HEPES. Lysis buffer (500µl, Mesa Photonics) was added to each well, incubated for 5 minutes at room temperature and 40µl of lysates added to duplicate wells of a 96 well white plate (Greiner). Peptides (40µl) in 2x assay buffer (100mM HEPES, 10mM DTT, pH 7.4) were added to the wells to achieve final concentrations of 2.5µM ACE2 peptide (FITC-YVADAPK-DNP) and 100 nM of Ubiquitin-AMC and 100nM ISG15-AMC. Fluorescence was read after 15 minutes and 1 hour in a fluorescence plate reader (Tecan Infinite, Morgan Hill, CA) with a 3 second shake between reads using ʎ_ex_=365 nm and ʎ_em_=465nm for AMC and ʎ_ex_=488nm and ʎ_em_=530nm for FITC.

### SARS-CoV-2 infection and protease activities

All experiments with SARS-CoV-2 propagation were performed in biosafety level 3 containment laboratories at the University of New Mexico Health Science Center (UNM HSC) with approved protocols.

#### Optimal infection time and MOI

VeroE6 cells (ATCC, Manassas, VA, USA, Cat# CRL-1586) were grown to 90% confluency (approx. 3× 10^5^ cells per well) in a 12-well plate with Dulbecco’s minimal essential medium (DMEM) supplemented with 10% fetal calf serum (FCS) and 1% penicillin/streptomycin. Cells were infected with 0.1 or 0.01 MOI of the SARS-CoV-2 isolate USA-WA1/2020 (BEI Resources, Manassas, VA, USA) in viral growth medium (MEM + 2.5% heat inactivated fetal calf serum) and incubated at 37°C in a humidified incubator for 2, 4, 8, and 24 hours. After each incubation time, cell-based protease assays were performed as described below.

#### Cell-based PLpro inhibition assays

The well-characterized PLpro inhibitor GRL0617 (R&D Systems) was diluted in MEM without fetal calf serum to achieve concentrations between 2µM-100µM and 100µl was added to near confluent VeroE6 cells in wells of a 24-well plate. Cells were incubated for 2 hours at 37°C in a humidified incubator, washed with 1x PBS, and 0.01 MOI of SARS-CoV-2 USA-WA1/2020 was added in viral growth medium (MEM + 2.5% heat inactivated fetal calf serum). Controls included medium with SARS-CoV-2 with and without DMSO (1% for a final concentration of 0.25%) and medium only with and without DMSO. Cells were incubated at 37°C in a humidified incubator for 24 hours. Cell-based protease assays were performed as described below.

#### Neutralization assays

Patient serum was diluted 1:40, 1:160, 1:640 and 1:1250 in viral growth media (MEM + 2.5% heat inactivated fetal calf serum) and 200µl was added to an equal volume of 0.01 MOI SARS-CoV-2 USA-WA1/2020 in viral growth medium for 2 hours. Controls were medium with SARS-CoV-2 without serum and medium without virus. The mixtures (400µl) were added to 90% confluent VeroE6 cells in 12-well plates and the cells were incubated at 37°C in a humidified incubator for 24 hours. Cell-based protease assays were performed as described below.

#### SARS-CoV-2 protease activity assays

After cells were incubated with virus for 6-24 hours, the media was removed from the wells and cells washed three times with 100 mM HEPES. Lysis buffer (100µl, Mesa Photonics) was added to each well, incubated for 5 minutes at room temperature and 25µl of lysates added to duplicate wells of a 96 well black plate (Corning). Peptides (25µl) in 2x assay buffer (100 mM HEPES, 10 mM DTT, pH 7.4) were added to the wells to achieve final concentrations of 2µM DEVD_2_-AMC and 50 nM Pun74-R110. Fluorescence was read immediately and every 15 minutes for 1 hour in a fluorescence plate reader (Synergy H1, Agilent) with a 3 second shake between reads using ʎ_ex_=365nm and ʎ_em_=465nm for AMC and ʎ_ex_=485nm and ʎ_em_=528nm for R110.

### Host cell lysis and SARS-CoV-2 inactivation by lysis buffer

All experiments determining the lysis of host cells and inactivation of SARS-CoV-2 using Mesa Photonics proprietary lysis buffer were performed in biosafety level 3 containment laboratories at MicroBiologics (MBL, St. Cloud MN) with approved protocols.

#### Host cell cytotoxicity testing

The ability of the lysis buffer to disrupt mammalian cell membranes was assessed by scoring the lysis buffer-caused morphological changes of VeroE6 cells and by measuring the effect of the lysis buffer on cell viability using the alamarBlue (Invitrogen) cell viability fluorescence staining reagent. Lysis buffer was used directly or filtered through Pierce detergent removal spin columns (PDRSC) according to the manufacturer’s recommendations (Thermo Fisher). 1:10 dilutions of lysis buffer with or without filtration by PDRSC and diluent (water) were prepared in viral growth media (VGM; 2% FBS in MEM with sodium pyruvate, non-essential amino acids, and antimycotic). Vero E6 cells (MBL) were grown in 6-well plates to 90% confluency in cell maintenance media (DMEM with 10% FBS, sodium pyruvate, and non-essential amino acids). The medium was replaced with 500µL of fresh VGM and then 100µl of lysis buffer dilutions or water were added. To mimic the binding step in plaque assays, the plates were incubated with shaking for 2 hours, the inoculates removed and replaced with 3mL fresh VGM. Cells were observed under an inverted microscope and then the plates were incubated at 37°C (5-6% CO_2_). After 7 days, the cell morphology was assessed by microscopy. For viability staining, VGM was removed from each well and replaced with 1mL of 10-fold diluted alamarBlue reagent prepared in VGM. After 4 hours incubation at 37°C, 100µL from each well were transferred to wells in a 96-well microtiter plate with flat and clear bottoms (Greiner, cat # 655801). Fluorescence signals were measured using a SpectraMax iD5 microtiter plate reader (Molecular Devices, Sunnyvale, CA) using excitation and emission wavelengths of ʎ_ex_=560nm and ʎ_em_= 590nm.

#### Virus inactivation testing using plaque assay

Viral inactivation was determined using SARS-CoV-2 stocks from 2 different lots of isolate USA-WA1/2020 virus (BEI NR-52281/Lot#: 70036318). For the virus only control group, viruses were prepared at 1:10 serial dilutions in VGM. For the experimental group, the viral stock was added to lysis buffer in a ratio of 1:9 and 1:10 serial dilutions prepared in VGM. The remaining infectivity in each sample dilution was determined using plaque assay in 1-day-old VeroE6 monolayer cultures (≥90 % confluency) prepared in 6-well cell culture plates. Briefly, cell maintenance medium was removed from the plated cells, and 500µL of VGM was added to each well. Then, 100µL of test dilutions were added to the corresponding wells. The plates were incubated under slow-speed shaking for 2 hours for virus-cell binding, then the inoculates were removed and 3mL of VGM contain 1.5% or 3% carboxy methyl cellulose (CMC; Teknova, Hollister, CA) were added followed by incubation at 37°C for 5-7 days in a CO_2_ Incubator. After incubation, the CMC overlay layer was aspirated and the cell monolayers were fixed by addition of 3 mL/well of 10% formaldehyde followed by a 30 min incubation. After removal of the formaldehyde, 2 mL of 0.1% Crystal Violet stain solution (Thermo Scientific) was added to each well for 10 minutes. The excess Crystal Violet was rinsed out with deionized (DI) water and the formed plaques were counted to calculate the retained infectivity titer.

### SARS-CoV-2 Protease Assays using Human Specimens

#### Ethical Approval

This project was approved by the University of New Mexico Health Sciences Center Human Research Protection Program. Review and approval were obtained from the UNM Institutional Review Board (IRB) under Study Numbers 22-373 and 22-038. All relevant ethical guidelines and regulations were followed, and study participants provided written informed consent. None of the subjects were taking antivirals and were directed to refrain from smoking, chewing tobacco, eating, or drinking 60 minutes prior to specimen collection.

#### SARS-CoV-2 PLpro activity assays in lysates from tongue scrape and saliva

The study entitled “A new, Simple On-Site Test for Detection of SARS Coronaviruses” compared the performance of the SARS-CoV-2 PLpro protease assay between lysates obtained from saliva and tongue scrape specimens.

#### Study population

14 male and 9 female subjects were enrolled between May 23, 2022 and June 23, 2022 after presenting to the Emergency Department with clinical signs of respiratory disease. COVID-19 was confirmed or ruled out by RT-PCR using a diagnostic standard of care test that had been approved under the FDA Emergency Use Authorization Framework (Cobas LIAT RT-PCR using nasopharyngeal swabs). 13 patients with positive and 10 patients with negative RT-PCR results were enrolled in the study.

Tongue scrape specimens were obtained by gentle and painless scraping of the tongue three times from back to front using a disposable tongue cleaner (Grin Oral Care, San Diego, CA). Tongue cleaners together with the adhering samples were placed in 50mL flat bottom sample collection tubes (Environmental Express), and 1mL 2x lysis buffer was added using a 1mL transfer pipette (Sigma Aldrich, St. Louis MO).

Saliva specimens were obtained by passive drooling through a straw. Raw saliva was transferred to a vial containing 200µL 2x lysis buffer using a 200µl Samco™ Exact Volume Transfer Pipette (Thermo Scientific).

Lysates were briefly vortexed and 200µl transferred to a Reaction Vial containing freeze dried assay components in a 0.5mL PCR Tube (Promega, Madison, WI) for final concentrations of 5mM DTT, 50mM HEPES, 20µM PLpro peptide Pun16-AMC, and 20µM caspase peptide DEVD_2_-R110. In addition, 200µl lysate was transferred to a separate vial (QuantiVial) containing the nucleic acid intercalating “Quanti dye”. Reaction Vials were inserted into a PCR tube adapter (Promega, E6101) and the fluorescence measured in a battery-operated fluorometer (Turner Designs, San Jose, CA) with 2 channels configured for dual fluorescence measurements: Channel A=UV LED, 365nm excitation filter, 440/15nm emission bandpass filter was used to measure fluorescence from the PLpro peptide Pun16 labeled with AMC. Channel B=Blue LED, 475nm excitation filter, 515/10nm emission bandpass filter was used to measure fluorescence from the internal control peptide DEVD_2_-R110. Fluorescence from the Reaction Vial was measured immediately (time 0) and every 30 minutes for 1 hour (time 60) in both channels. The fluorescence from the QuantiVial was measured once after 1 hour in Channel B. Values were recorded manually and the difference between relative fluorescence units (RFU) measured at time 60 and time 0 calculated (delta RFU). The delta RFU were normalized for the amount of material collected using the RFU measured in the QuantiVial according to the formula (Delta RFU protease/RFU QuantiVial)*100.

#### SARS-CoV-2 PLpro / MPro assays in lysates from saliva specimens

A second study entitled “A new, Simple On-Site Test for Detection of SARS Coronaviruses in Saliva” compared the performance on saliva that was split for the SARS-CoV-2 protease assays and for RT-PCR on purified RNA.

#### Study population

9 male and 13 female subjects were enrolled between June 6, 2023, and September 27, 2023, who presented to the Emergency Department with symptoms consistent with COVID-19. A positive or negative RT-PCR diagnosis was made using standard of care Cobas LIAT RT-PCR on nasal mid-turbinate (NMT) swabs. None of the subjects were taking antivirals and were directed to refrain from smoking, chewing tobacco, eating, or drinking 60 minutes prior to specimen collection.

Saliva specimens (500µl) were collected using a SpeciMAX^TM^ Saliva Collection kit (Thermo Scientific, Waltham, MA). For the protease tests, 200µl lysate was added to vials containing 200µl 2X Lysis buffer (Mesa Photonics). In this study, SARS-CoV-2 PLpro and MPro protease assays were performed in 384-well plates and the fluorescence monitored in a fluorescence plate reader. Lysate (10 µl) was added to triplicate wells and reactions started by addition of 10µl reaction mixtures. Reaction mixtures contained A) the MPro-R110 peptide and B) the PLpro peptide Pun16-R110 to yield final concentrations of 2µM peptide in 5mM DTT and 50mM HEPES, pH 7.4); C) a 2.5x dilution of Quanti dye was added to separate triplicate wells containing lysate. Fluorescence was monitored every 2 minutes in a fluorescence plate reader for 30 minutes using ʎ_ex_=485nm and ʎ_em_=528nm at 30 °C.

<u>Data analysis.</u> For MPro assays, the delta RFU was calculated from the difference in relative fluorescence units (RFU) measured at time 30 and time 0 and for PLpro assays, the delta RFU was calculated from the difference in RFU measured at time 30 and time 10. The coefficient of variation (CV) was calculated using the formula: (standard deviation / mean of RFU from triplicate values)*100. The Delta RFU were normalized for the amount of sample collected by calculating the ratio between the RFU from the protease assays and Quanti dye using the formula (Delta RFU Protease assay/RFU Quanti dye)*100. The ratio of CV was calculated using the formula *Ratio CV* = √(*CVPLpro*^2^ + *CVQV*^2^). Pearson correlations were determined using GraphPad Prism software version 10.2.0 (Boston, MA). The limit of detection (LoD) of MPro and PLpro activities was determined by averaging the delta RFU and ratios from 5 representative experiments + 3x stdev. The specimen LoD (via SYBR dye) was determined by serially diluting a positive and a negative specimen in lysis buffer and measuring the delta RFU from MPro and PLpro peptide cleavage and the RFU from SYBR dye. The point of the Y intercept (corresponding to lysate dilution) for the positive and negative specimen was calculated as follows: x = b2-b1/m1-m2, where b=intercept and m=slope. The values were interpolated to the RFU from the SYBR dye linear regression curve using the formula y=mx+b.

#### RNA Isolation

Saliva (200µl) from the same specimen used for the protease assays was transferred to a vial containing an equal volume of RNA Shield (Zymo Research, Irvine, CA). RNA was isolated using the Zymo Research Quick-DNA/RNA Viral Kit according to manufacturer’s recommendations. SARS-CoV-2 LAMP RT-PCR and TaqPath One step PCR were performed as described below.

#### LAMP SARS-CoV-2 RT-PCR

LAMP PCR was performed using the SARS-CoV-2 Rapid Colorimetric LAMP Assay Kit from New England Biolabs (Ipswich, MA) according to the manufacturer’s instructions with the supplied primers that target the SARS-CoV-2 nucleocapsid (N) gene, the envelope (E) gene, and the internal control actin. LAMP Fluorescent Dye (NEB) was added at a 1x final concentration to enable real-time fluorescence monitoring. The reactions were prepared in Fast Reaction Tubes with Cap (Thermo Fisher), overlayed with 20µl mineral oil, and incubated for 30 minutes at room temperature. Tubes were placed in an AXYGEN PCR Tube Storage Rack (#80086-082, Corning, NY) and the rack inserted into a Synergy H1 fluorescent plate reader using the following plate settings: Biotek PCR adapter 2 cm plate, bottom read, read height 7 mm. Fluorescence was measured every 2 minutes for 60 minutes at 65 °C in kinetic mode using excitation and emission wavelengths of ʎ_ex_=485nm and ʎ_em_=528nm.

#### Endpoint SARS-CoV-2 RT-PCR

The TaqPath™ 1-Step Multiplex Master Mix (no ROX, Thermo Scientific) was used to amplify 2µl RNA according to the manufacturer’s instruction for FAST cycling. Primers for SARS-CoV-2 N1, N2, and RNase P genes were purchased from Integrated DNA Technologies (Coralville, IA). The sequences correspond to those supplied with the 2019-nCov CDC EUA Kit.^28^

Reactions (20µl) were prepared in 0.2mL PCR tubes, overlayed with mineral oil, and RT-PCR performed in a MiniAmp Thermocycler (Thermo Fisher) using thermal cycling conditions of 2 minutes at 25 °C (UNG incubation), 10 minutes at 53 °C (reverse transcription), 2 minutes at 95 °C (polymerase activation) and 40 cycles of 3 seconds at 95 °C and 30 seconds at 60 °C.

1.5µl Safestain 6x DNA Loading Dye (Lambda Biotech, St. Louis, MO) was added to 8µl PCR products and 10µl (0.8µg) 100 bp DNA Ladder (Lamda Biotech) or to 1µg 50 bp DNA Ladder (New England Biolabs). Samples (8µl) were loaded into wells of a 2% agarose gel and electrophoresis performed for 1-2 hours at 40 Volts (∼1 Watt) in Tris Borate EDTA (TBE) buffer. Peak intensities of the bands were quantified using ImageJ and the percentage of SARS amplicon intensities calculated relative to the intensities of the RNase P amplicons.^29^

## RESULTS

### Detection of mono-ubiquitin and ISG15 cleavage in lysates from human-infecting coronaviruses

Papain-like proteases are expressed by other human coronaviruses (hCoV)—SARS-1, MERS, NL63, OC43, HKU1 and 229E—that also cleave the LRGG sequence in reporter substrates. To determine whether the PLpro assay was sensitive enough to detect papain-like protease activities in cell-based assays, we first tested human coronaviruses that could be grown in a biosafety level 2 laboratory. PLPs from both non-SARS-CoV-2 human-infecting coronaviruses and SARS-CoV-2 PLpro reportedly have deubiquitinating (DUB) and deISGylating activities that, in the case of SARS-CoV-2 PLpro, have an enzymatic efficiency over 2,000 times higher for a ISG15-AMC substrate relative to the tetra peptide-AMC in a biochemical assay.^30,31^ Accordingly, in our *in vivo* PLpro assay, we were able to detect cleavage of ISG15-AMC in lysates from approximately 24,000 human MRC-5 and HCT8 cells infected with the human coronaviruses 229E and OC43, respectively. Mono ubiquitin-AMC cleavage was detectable after 15 minutes in lysates from LLC-MK2 cells that were infected with NL63 (Figure 2, Supplementary Information) but we were unable to observe stable signals from tetrapeptide cleavage in any of the lysates tested.

**Figure 2.**
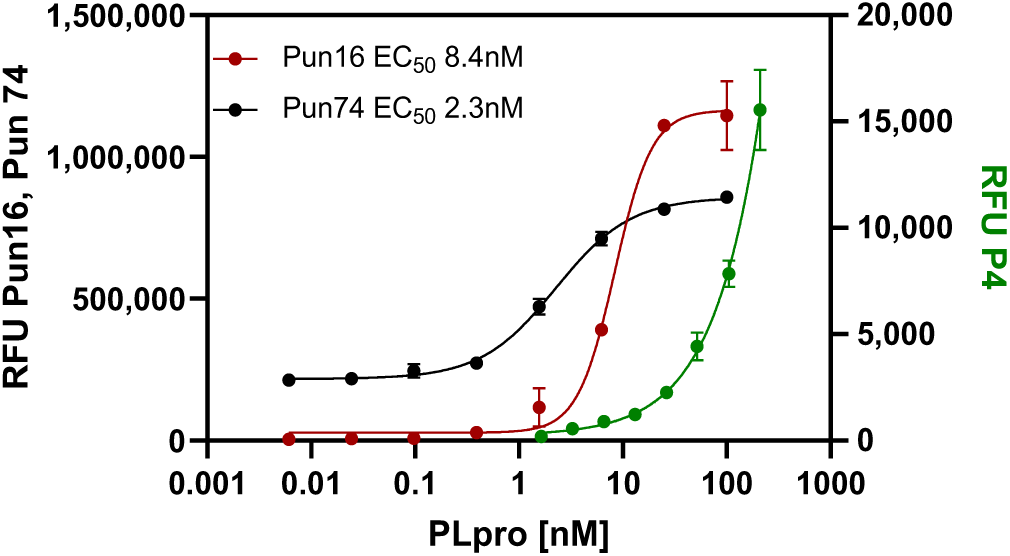
Effect of PLpro peptide length on detection sensitivity. A serial dilution of recombinant PLpro was prepared in assay buffer and added to duplicate wells of a 384-well plate. Peptides in assay buffer were added and the plate incubated for 60 minutes at room temperature. The final concentration of Pun74-R110 was 300nM and 30µM for P16-AMC and P4-AMC. Fluorescence was monitored using dual wavelength monitoring for R110 (ʎ_ex_=485 nm and ʎ_em_=528 nm) and AMC (ʎ_ex_=365 nm and ʎ_em_=465 nm). Error bars represent standard deviations of the mean relative fluorescent units (RFU) from duplicate wells.

**Figure 3.**
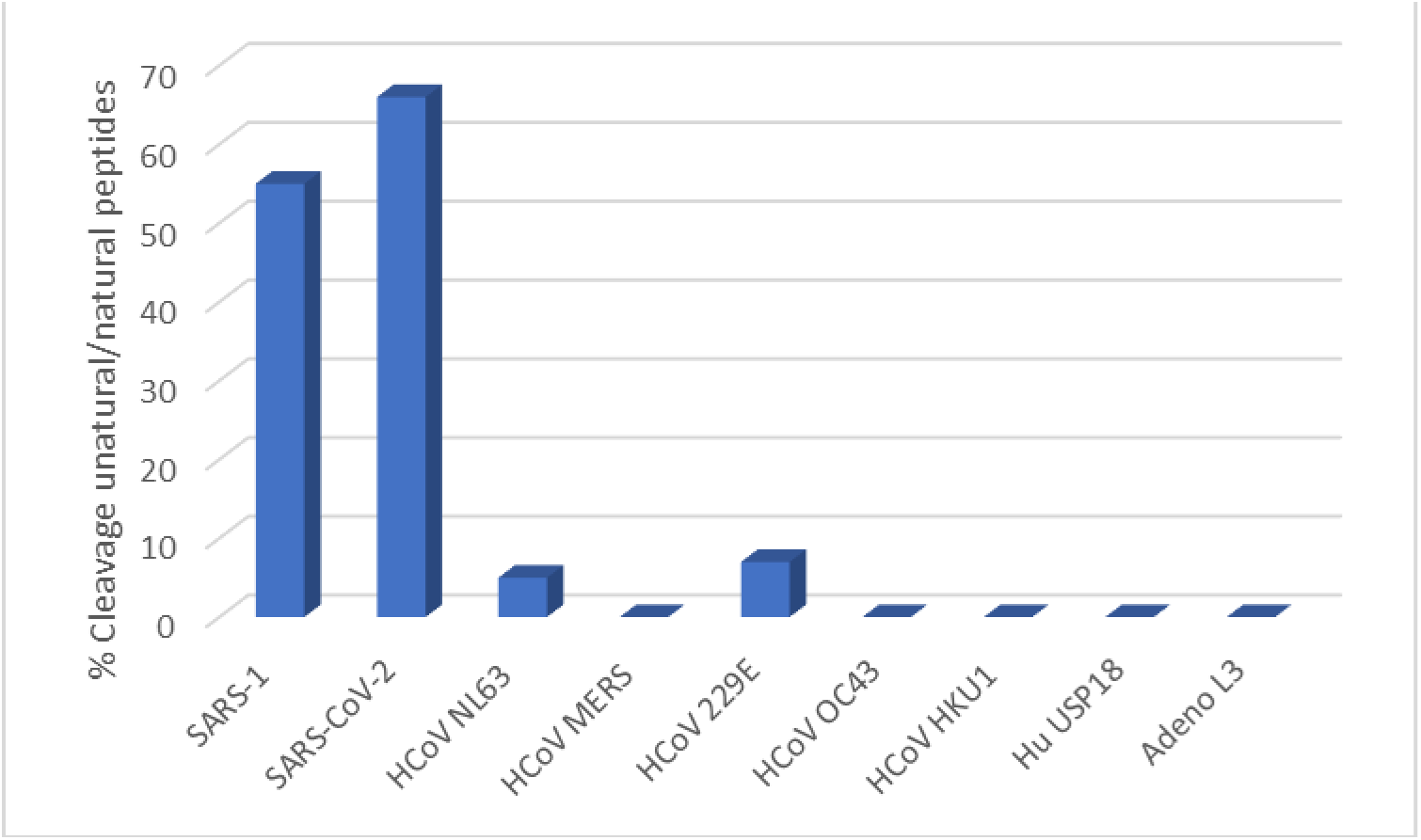
Cleavage of PLpro peptide Pun16-AMC by various papain-like proteases. The fluorescence from the cleavage of PLpro peptide Pun16-AMC with unnatural amino acids in the cleavage site and from the cleavage of positive control substrates with natural amino acids was measured. The concentrations of enzymes were 100nM (SARS-1, SARS-CoV-2 PLPro), 250nM (Adenovirus L3), 1200nM (MERS PLPro, 229E PLP2, HKU1 PLP2, USP18), and 1760nM (OC43 PLP2). The concentration of positive control substrate P16 was 25µM for PLPs from SARS-1, SARS-CoV-2, NL63, MERS, 229E, and 10µM Ubiquitin-AMC for PLPs from OC43 and HKU1, 0.5µM ISG15-R110 for USP18 and 60µM P4 for adenovirus L3 protease. The reaction progress was monitored for 60 minutes at room temperature and the differences in RFU between time 60 and time 0 calculated (delta RFU). The percent cleavage activity of peptide Pun16-AMC was calculated relative to the delta RFU of the control substrates ((delta RFU_control_/deltaRFU_Pun16_)*100).

### Peptides with improved sensitivity and specificity of detection

In a next step, we aimed to combine the observed sensitivity of detection afforded by using peptides containing sequences of ISG15 with specificity of detection required for a SARS-CoV-2 diagnostic test. Specificity was conferred by incorporating unnatural amino acids into the PLpro cleavage site following Rut et al., who showed that PLpro from SARS-1 and SARS-CoV-2 have unique preferences for amino acids in the LXGG cleavage sequence (where L= leucine at position 4, X=any amino acid at position 3, G=glycine at position 2, and G= glycine at position 1). They found that only glycine is accepted at positions 1 and 2, a broad range of amino acids are accepted at position 3, and hydrophobic residues only are accepted at positions 4.^32^ We therefore synthesized peptides containing 16 or 74 amino acids from ISG15 and unnatural amino acids in the cleavage site (Pun16 and Pun74).

The increased performance of the longer peptide relative to the tetrapeptide P4-AMC was estimated in a biochemical assay by determining the concentration of recombinant PLpro required to produce a half maximum signal (EC_50_). The tetrapeptide P4-AMC did not saturate the enzyme at the highest concentration tested and the EC_50_ was estimated as half the signal after 1 hour of incubation (EC_50_=100 nM). This resulted in an estimated 12 and 43.4-fold increase in detection performance for a 16-mer peptide (Pun16-AMC; EC_50_= 8.4nM) and 74-mer peptide (Pun74-R110; EC_50_= 2.3nM) (Figure 2).

Next, we tested cleavage of the ISG15-based peptide Pun16-AMC that contained unnatural amino acids at positions 4 and 3 by recombinant PLPs from other human pathogens known to have deISGylating activities. We show that Pun16-AMC was significantly cleaved by recombinant PLpros from SARS-1 and SARS-CoV-2 but not by recombinant PLpro/PLPs from the other five human coronaviruses, by the human deISGylating enzyme USP18, or Adenovirus L3. We verified that all recombinant PLPs were catalytically active by measuring the RFU resulting from cleavage of positive control substrates that contained only natural amino acids in the cleavage sites. The positive control peptide P16-AMC has the same amino acid sequence as Pun16-AMC but with the natural amino acids leucine (L) and arginine (R) in the cleavage sequence. P16-AMC was cleaved by recombinant papain-like proteases from SARS-1, SARS-CoV-2, NL63, MERS, and 229E but not by PLPs from OC43 and HKU1; for these enzymes, ubiquitin-AMC was used as the positive control substrate instead. The positive control substrate for USP18 was human ISG15-R110, and the positive control substrate for the adenovirus L3 protease was the tetrapeptide P4-AMC.

The study with recombinant enzyme showed that the PLpro peptide Pun16-AMC with unnatural amino acids in the cleavage site was strongly cleaved by PLpros from SARS-1 and SARS-CoV-2 (53.6% and 65.6% relative to P16-AMC, respectively), minimally cleavage by PLPs from HCoV NL63 and 229E (5.8% and 7.7% relative to P16-AMC, respectively), and not detectably cleaved by the other proteases tested. The lack of cross-reactive cleavages supported the expectation that cleavage activities measured in crude lysates would be predominantly from SARS-CoV-2 PLpro and not from proteases of other human coronaviruses or endogenous proteases that may be present in lysates.

### Detection of PLpro and caspase activities in crude lysates from SARS-CoV-2 infected VeroE6 cells

Next, we tested whether the PLpro Pun peptides, which were designed and optimized for cleavage by SARS-CoV-2 PLpro *in vitro*, could produce fluorescent signals in crude lysates from SARS-CoV-2 infected VeroE6 cells that could be discriminated from background fluorescence produced by potential host cell protease cleavage. When lysates from 3×10^5^ VeroE6 cells were tested that were infected with a multiplicity of infection (MOI) of 0.1, we observed strong fluorescent signal increases predominantly in lysates from infected cells and not in those from uninfected cells that were run in parallel as a control (Figure 4, left). After 60 minutes of incubation with Pun74-R110, the ratio of RFU at 60 minutes/RFU at time 0 (Signal to Background) from infected cell lysates was 16 while the S/B from uninfected cell lysates was only 1.8. This demonstrates that 90% of the measured fluorescence resulted from cleavage of the PLpro peptide Pun74-R110 by PLpro rather from endogenous proteases present in the lysate.

**Figure 4.**
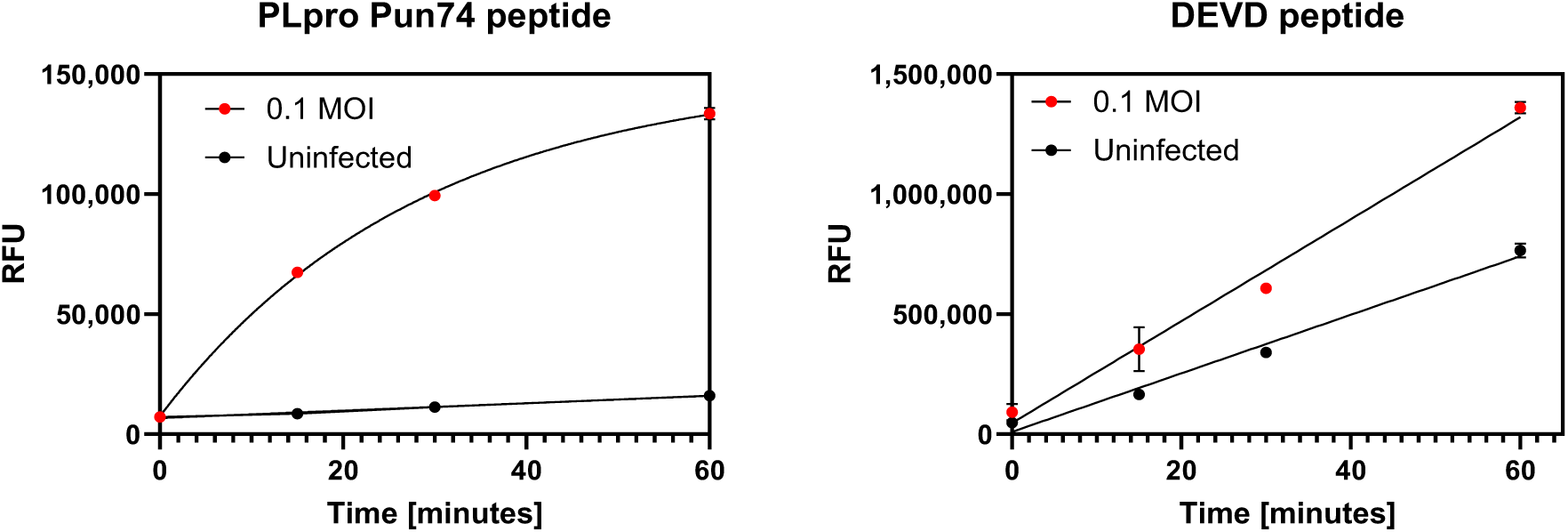
Fluorescence increases from cleavage of PLpro and DEVD peptides by SARS-CoV-2 PLpro and caspases in crude lysates from SARS-CoV-2 infected VeroE6 cells. VeroE6 cells (3×10^5^) were infected with 0.1 MOI SARS-CoV-2 for 12 hours and lysates prepared from infected (red traces) and uninfected (black traces) cells. Lysates (25µl) were added to wells of a 96-well plate and PLpro peptide Pun74-R110 (50nM) or DEVD_2_-R110 (2µM) in 25µl 2x assay buffer added. The increase in relative fluorescence units (RFU) in x2 replicates was monitored immediately and after every 15 minutes for 60 minutes at room temperature in a fluorescence plate reader using excitation and emission wavelengths of ʎ_ex_=485nm and ʎ_em_=528nm. RFU were plotted using GraphPad Prism software. Some error bars are within the symbols. Pun74 substrate depletion in lysates from infected cells resulted in non-linear fluorescence increases after approximately 15 minutes. The results were reproducible in five separate experiments.

We also measured the fluorescence resulting from the cleavage of the peptide with the sequence DEVD, which is recognized by caspases, and determined that this peptide can be used as an internal processing control to confirm that lysates were present in sufficient quantity and quality for the assay to perform. As expected, infected cells demonstrated higher cleavage of the DEVD peptide than uninfected cells as the cleaving enzymes, caspases, are upregulated in pathogen-infected cells preparing to undergo apoptosis.^33^

### Time course of PLpro activity in lysates from SARS-CoV-2 infected VeroE6 cells

To determine the optimal incubation time and multiplicity of infection for detecting SARS-CoV-2 PLpro activity, we incubated SARS-CoV-2 at MOIs of 0.1 and 0.01 with VeroE6 cells and prepared lysates after various incubation times. The PLpro peptide was added to the lysates and the S/B was calculated between the start of the reaction and after 60 minutes of incubation (Figure 5). We found that lysates infected with an MOI of 0.1 produced a S/B of 12 after 6 hours of incubation, which was approximately 4 times greater than the S/B of 6 that was obtained in lysates from uninfected cells. These results correspond to the reported detection of intracellular SARS-CoV-2 virions ∼10 hours post infection which require PLpro activity for assembly.^15^ In contrast, lysates from cells that were infected with an MOI of 0.01 required an incubation time of approximately 24 hours to achieve a noticeable difference relative to uninfected cells. Although the experiment was performed only once, the results suggest that assays can be completed within 1 working day from infection to result using an MOI of 0.1.

**Figure 5.**
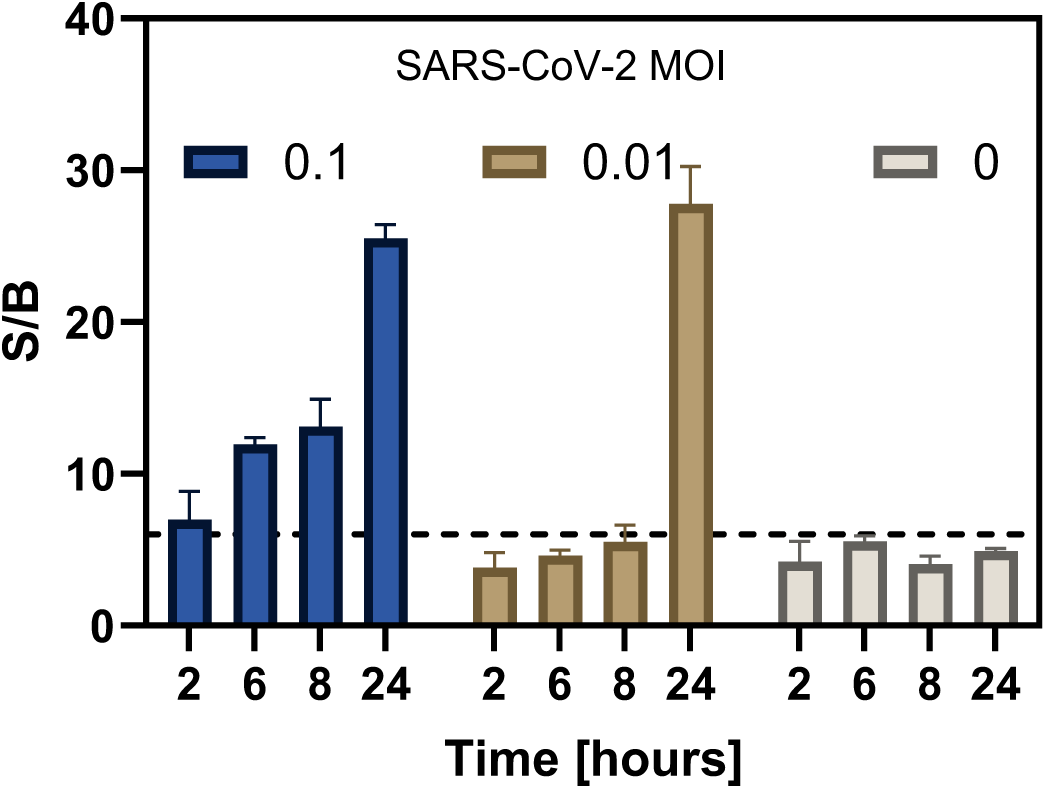
Time course of fluorescence increases in lysates from VeroE6 cells infected with SARS-CoV-2. VeroE6 cells (∼300,000 cells per well) were plated in a 12-well plate. 4 wells were infected with a MOI of 0.1 or 0.01. Control cells were not infected and treated with medium. Lysis buffer (100µl) was added to washed monolayers and 25µl of lysates (corresponding to 75,000 cells) from each time point were combined with the PLpro substrate Pun74-R110 in wells of a 96-well plate. The increases in relative fluorescence units (RFU) were measured in x2 replicates immediately and after 60 minutes in a fluorescence plate reader using excitation and emission wavelengths of ʎ_ex_=485nm and ʎ_em_=528nm. The ratio of RFU between the 60-minute read and the initial read was calculated and plotted as Signal to Background (S/B) using GraphPad Prism software. A S/B of 6 was used as a cutoff and is indicated by a dotted line.

### Measurement of PLpro inhibition in crude lysates from SARS-CoV-2 infected VeroE6 cells

The excellent performance of the PLpro assay in crude lysates from VeroE6-infected cells encouraged us to test whether the platform has potential for use as a screening tool to identify PLpro inhibitors. Feasibility was demonstrated using the well characterized PLpro inhibitor GRL0617.^34^ For the inhibition assay, various concentrations of inhibitor were added to VeroE6 cells and incubated for 2 hours. SARS-CoV-2 was added at an MOI of 0.01 for 24 hours, after which protease activities were measured as described. The results shown in Figure 6A demonstrate that a concentration of 100µM of the inhibitor reduced the fluorescence to nearly the background fluorescence of the wells containing uninfected lysates (medium only or medium + 0.25% DMSO without virus), while lower concentrations of inhibitor caused only a slight decrease in fluorescence relative to the controls (medium, medium + virus, and medium + DMSO no virus).

**Figure 6.**
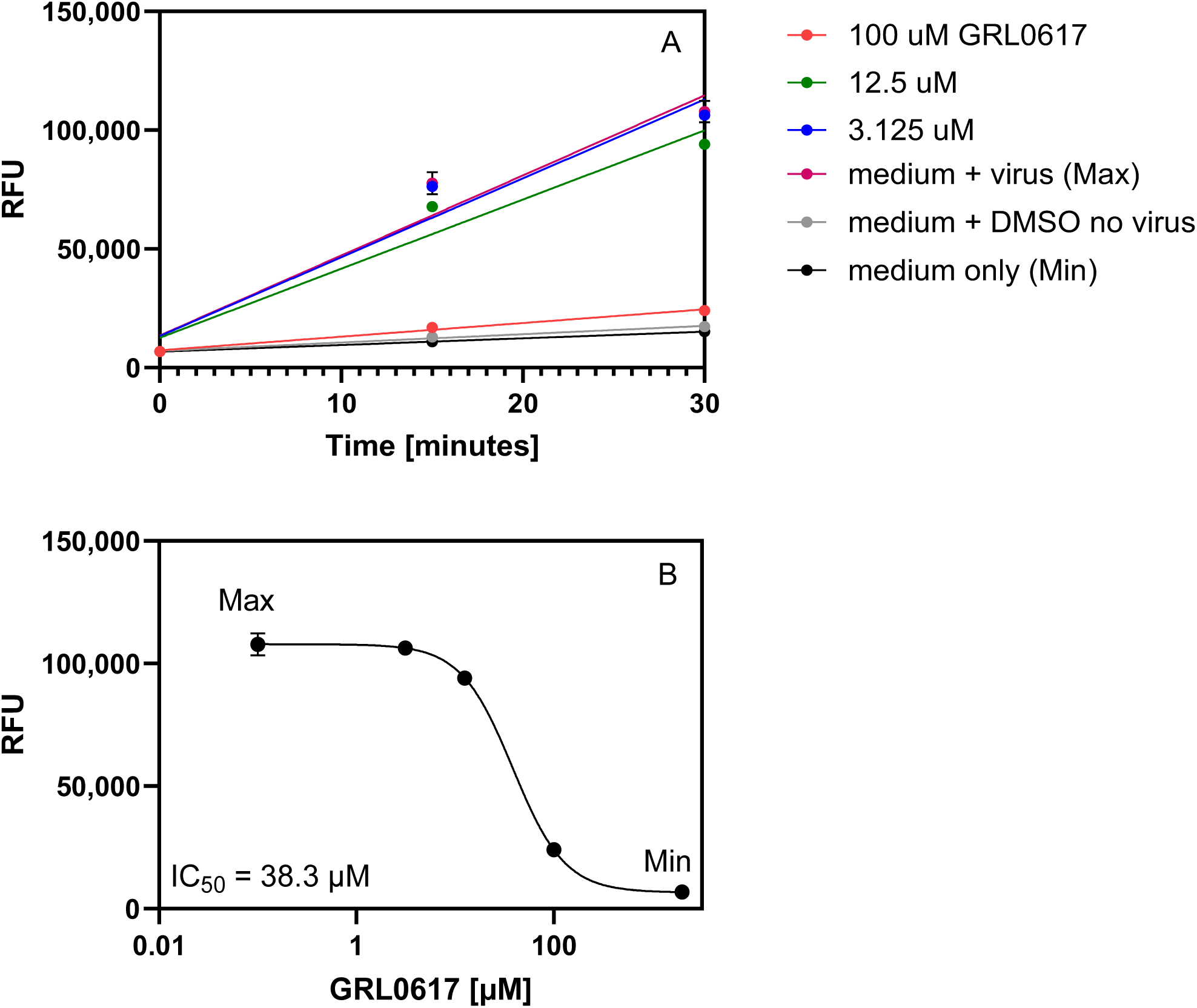
Fluorescence from PLpro peptide cleavage in lysates from VeroE6 cells infected with GRL0617 pretreated SARS-CoV-2. VeroE6 cells were seeded in a 24-well plate and incubated at 37 °C for 12 hours. Various dilutions of the inhibitor GRL0617 were prepared in MEM without FCS, added to the cells, and incubated for 2 hours at 37 °C. Control cells were uninfected and infected cells treated with medium only and uninfected cells treated with medium plus DMSO. SARS-CoV-2 (0.01 MOI) was added for 24 hours at 37 °C and lysates prepared. PLpro substrate Pun74 was added in 2x assay buffer and 25µl added to wells in a 96-well plate for a final concentration of 50nM. **A)** The increase in relative fluorescence units (RFU) from Pun74 cleavage in x2 replicates was monitored every 15 minutes for 30 minutes at room temperature in a fluorescence plate reader using excitation and emission wavelengths of ʎ_ex_=485nm and ʎ_em_=528nm. **B)** The mean RFU after 30 minutes in lysates from cells pretreated with GRL0617 and Max (medium plus virus) and Min (medium without virus) values were plotted and the IC_50_ calculated using GraphPad Prism, nonlinear regression, variable slope. Some error bars are within the symbols.

The tentative IC_50_ value of 38.3µM obtained for GRL0617 in our hands was within one order of magnitude of the EC_50_=21 ± 2 μM reported in a virus proliferation assay, demonstrating successful inhibition. Although this experiment was performed only once, the results suggest that the assay can be used for rapid screening of PLpro inhibitors against endogenous PLpro.^34^

### Detection of Serum Neutralizing Activities using SARS-CoV-2 PLpro as a marker of infection

A neutralization assay was performed to determine whether PLpro activity can be used as a marker of infection and determine the neutralizing activity of anti-SARS-CoV-2 antibodies. Building on the information gained from the time point experiment (Figure 5), we conducted a neutralization assay in which a MOI of 0.01 SARS-CoV-2 was combined for 2 hours with various dilutions of anti-SARS-CoV-2 positive serum. This mixture was then added to VeroE6 cells and lysates were prepared after 24 hours of incubation. The cell-based protease assay was performed as described and the delta RFU calculated after 30 minutes. Lysates from cells that were challenged with virus pretreated with serum dilution from 1:40, 1:160, and 1:640 produced fluorescence that was not distinguishable from the delta RFU obtained from uninfected control lysates (Figure 7, Medium), demonstrating complete neutralization. In contrast, lysates from cells in which SARS-CoV-2 had been incubated with the serum dilution of 1:2,560 produced delta RFU that was 33% lower than the delta RFU from untreated virus (Medium + virus), indicating that infection, albeit at a lower level, had occurred. If reproducible, these findings from a single experiment suggest that PLpro cleavage activity is a useful biomarker for rapidly determining antibody neutralization activity.

**Figure 7.**
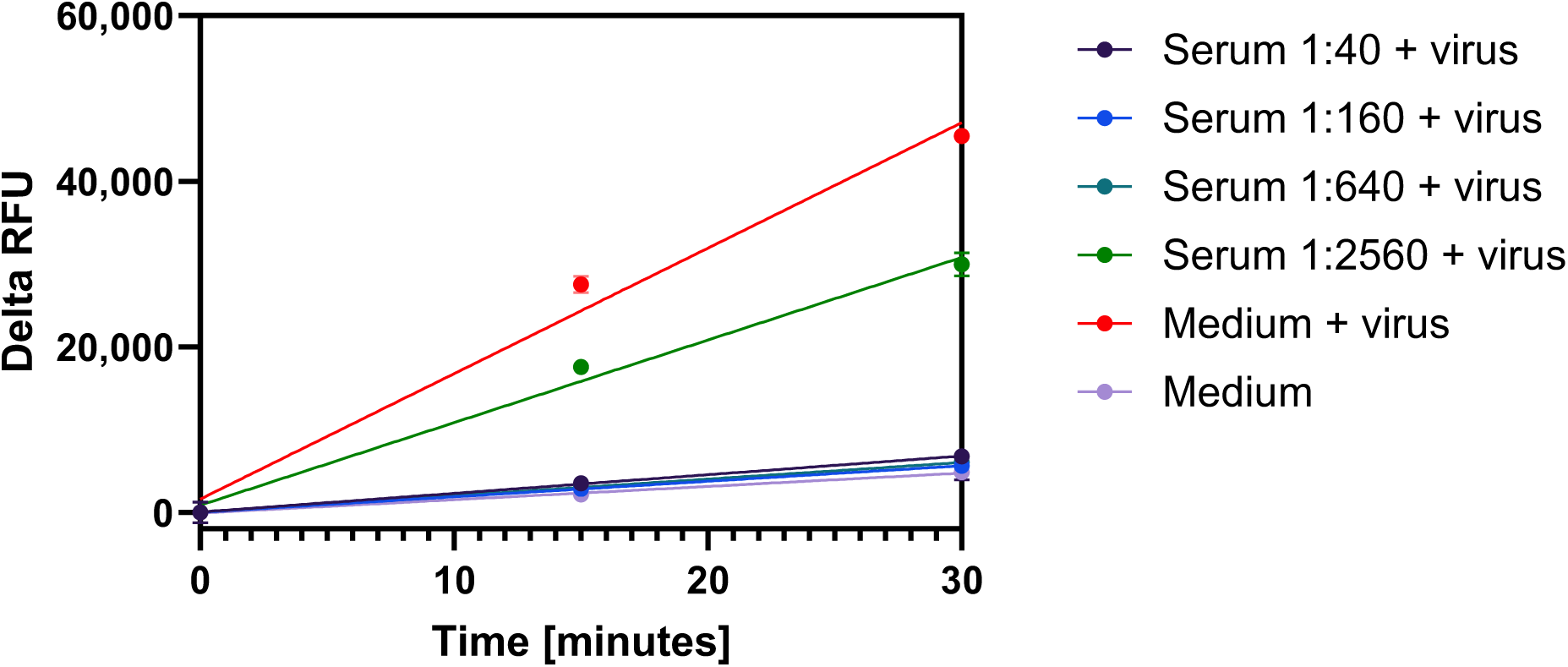
Fluorescence from PLpro peptide cleavage in lysates from VeroE6 cells infected with serum pretreated SARS-CoV-2. VeroE6 cells were seeded in a 12-well plate and incubated at 37 °C for 12 hours. Various dilutions of serum were prepared in viral growth medium and combined with equal volumes of 0.01 MOI of SARS-CoV-2 for 2 hours at 37 °C. The mixtures were added to the confluent VeroE6 cells for 24 hours at 37 °C. Control cells were uninfected cells treated with medium or infected cells treated with media only. Cells were washed, lysates prepared and transferred (25µl) to duplicate wells of a 96-well plate. PLpro substrate Pun74-R110 was added in 2x assay buffer for a final concentration of 50nM. The increase in relative fluorescence units (RFU) from peptide cleavage was monitored every 15 minutes for 30 minutes at room temperature in a fluorescence plate reader using excitation and emission wavelengths of ʎ_ex_=485nm and ʎ_em_=528nm. The difference in RFU between the 30-minute read and the initial read was calculated and plotted as Delta RFU using GraphPad Prism software.

### Host cell lysis and SARS-CoV-2 inactivation by Mesa Photonics lysis buffer

Handling of infectious SARS-CoV-2 is currently restricted to high-containment laboratories, but specimens can be handled at a lower containment level if potential virus is inactivated. The lysis buffer used for all experiments performed here is a proprietary mixture of detergents that do not interfere with the activities of the enzymes tested but is expected to disrupt the lipid membranes of host cells and virus, thus inactivating SARS-CoV-2. Complete host cell lysis was observed by microscopy when VeroE6 cells were treated with a 0.33x concentration of lysis buffer for 5 minutes, whereas 0.033x dilutions did not cause host cell lysis (Figure 3 Supplementary Information). The results were quantified by alamarBlue staining, where lysis buffer treated cells exhibited a 2-log reduction in fluorescence signal relative to the control groups (Figure 4, Supplementary Information).

Further, complete inactivation of 2 different lots of the SARS-CoV-2 WA strain (V00193 and V00282) was observed by plaque assay following a 5-minute incubation with lysis buffer, even at a 0.033x concentration, thus confirming that lysates containing a 1x dilution of lysis buffer can be safely handled during specimen testing (Figure 5, Supplementary data).

### Detection of SARS-CoV-2 PLpro activity in patient saliva and tongue scrape specimens in Point of Care Testing (POCT)

Following the successful demonstration of specific and sensitive cleavage of the PLpro Pun74 peptide in lysates from VeroE6 cells infected with SARS-CoV-2, we aimed to evaluate the performance of the test in lysates from human specimens. We first compared human tongue scrape and saliva specimens, given the reported abundance of epithelial cells on the tongue and in other tissues of the oral cavity that express the ACE2 receptor through which SARS-CoV-2 enters the cell.^35–38^ Saliva contains between 2-4 million epithelial cells/mL that are shed into saliva.^36^ Having been able to detect protease activities in lysates from 75,000 VeroE6 cells (Figure 5) we hypothesized that the assay would have sufficient sensitivity to detect PLpro and caspase cleavage activities in 100µl saliva, estimated to contain 200,000 to 400,000 potentially infected epithelial cells.^36^

Our objective was to compare the performance of the test in lysates derived from tongue scrape specimens and saliva, to determine if one type of specimen had an advantage over the other. We collected paired specimens from a total of 23 subjects who had given their informed consent and had been instructed to abstain from eating, drinking, or using tobacco products for 60 minutes prior to specimen collection and had not taken antivirals. The specimens were inactivated by addition of lysis buffer.

Testing was performed by minimally trained operators in a setting closely resembling a Point of Care Testing (POCT) site, using a portable battery-operated fluorometer and vials containing freeze-dried reaction components that did not require refrigeration or special storage conditions. The two-channel fluorometer was configured specifically for our test to enable us to measure the fluorescence produced by the PLpro peptide Pun16 (labeled with AMC) and the internal processing control DEVD peptide (labeled with Rh110) in a single sample vial. Fluorescence signals were normalized for the amount of lysate present by measuring the fluorescence from the nucleic acid intercalating dye (QuantiVial) in a separate specimen vial. The steps involved in the measurement process are illustrated in Figure 8.

**Figure 8.**
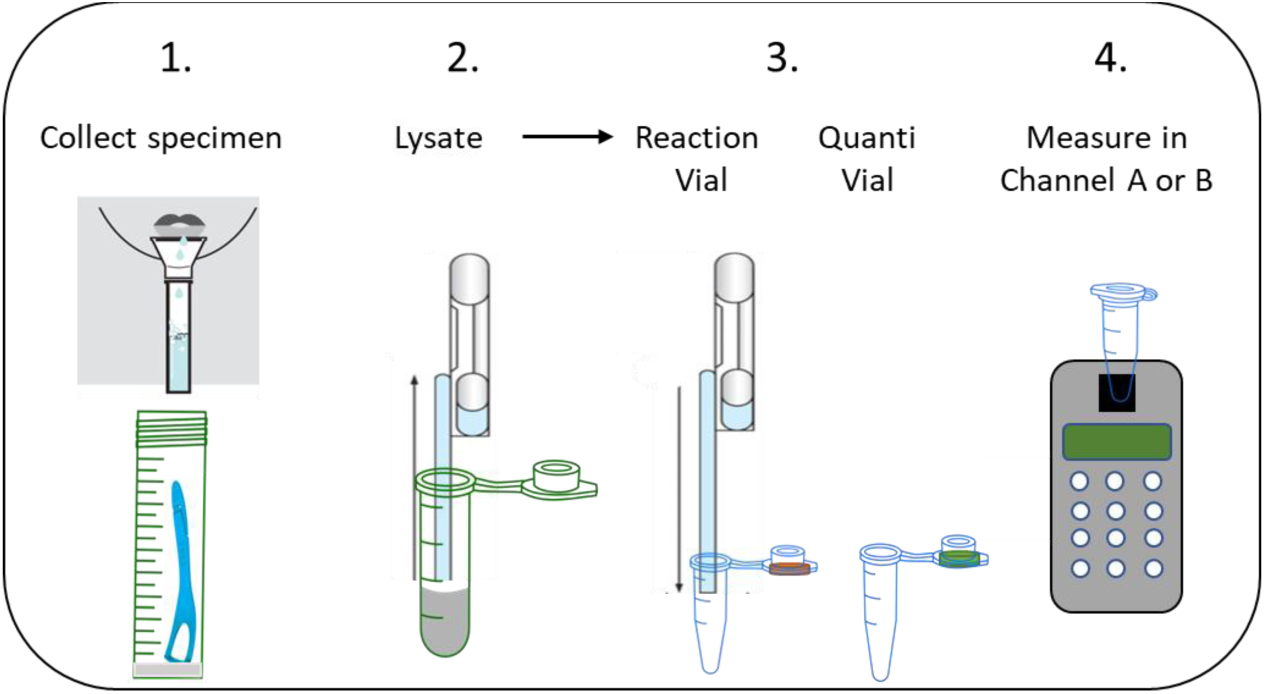
Procedure for Measuring SARS-CoV-2 PLpro Activity in a Point of Care Testing (POCT) setting. **(1)** Saliva was collected either through passive drooling using a straw or with a saliva collection device. A volume of 200µl of raw saliva was then transferred to a vial containing 200µl of 2x Lysis Buffer which immediately lyses cells and inactivates SARS-CoV-2. Painless tongue scrape specimens were collected using a commercial disposable tongue scraper and deposited into a tube containing 1 ml 2x Lysis Buffer. **(2)** The lysates were transferred using a disposable Exact Pipette and **(3)** 200µl was added to a Reaction Vial that contained lyophilized mixtures of reporter peptides for SARS-CoV-2 PLpro and caspases in assay buffer. An additional 200µl of the lysate was transferred to a separate vial containing lyophilized DNA intercalating dye (Quanti Vial) that was used to normalize the fluorescence signal to the amount of sample present. **(4)** The reaction mixtures were briefly mixed, and the vials inserted into the port of a battery-operated fluorometer that was configured to read fluorescence with excitation and emission wavelengths of ʎ_ex_=360nm and ʎ_em_=460nm (Channel A for PLpro peptide Pun16) or ʎ_ex_=490nm and ʎ_em_=520nm (Channel B for DEVD internal control peptide). Fluorescence from the Reaction Vial was monitored immediately and every 30 minutes for 1 hour and the values recorded manually. At the end of the experiment, the fluorescence from the Quanti Vial was measured in Channel B. The normalized delta RFU were calculated using the formula ((RFU_t=60_ - RFU_t=0_)/RFU QuantiVial)*100.

Lysates from tongue scrapes and saliva specimens were tested in parallel. Examples of a positive result, a negative result, and an inconclusive result are shown in Figure 9.

**Figure 9.**
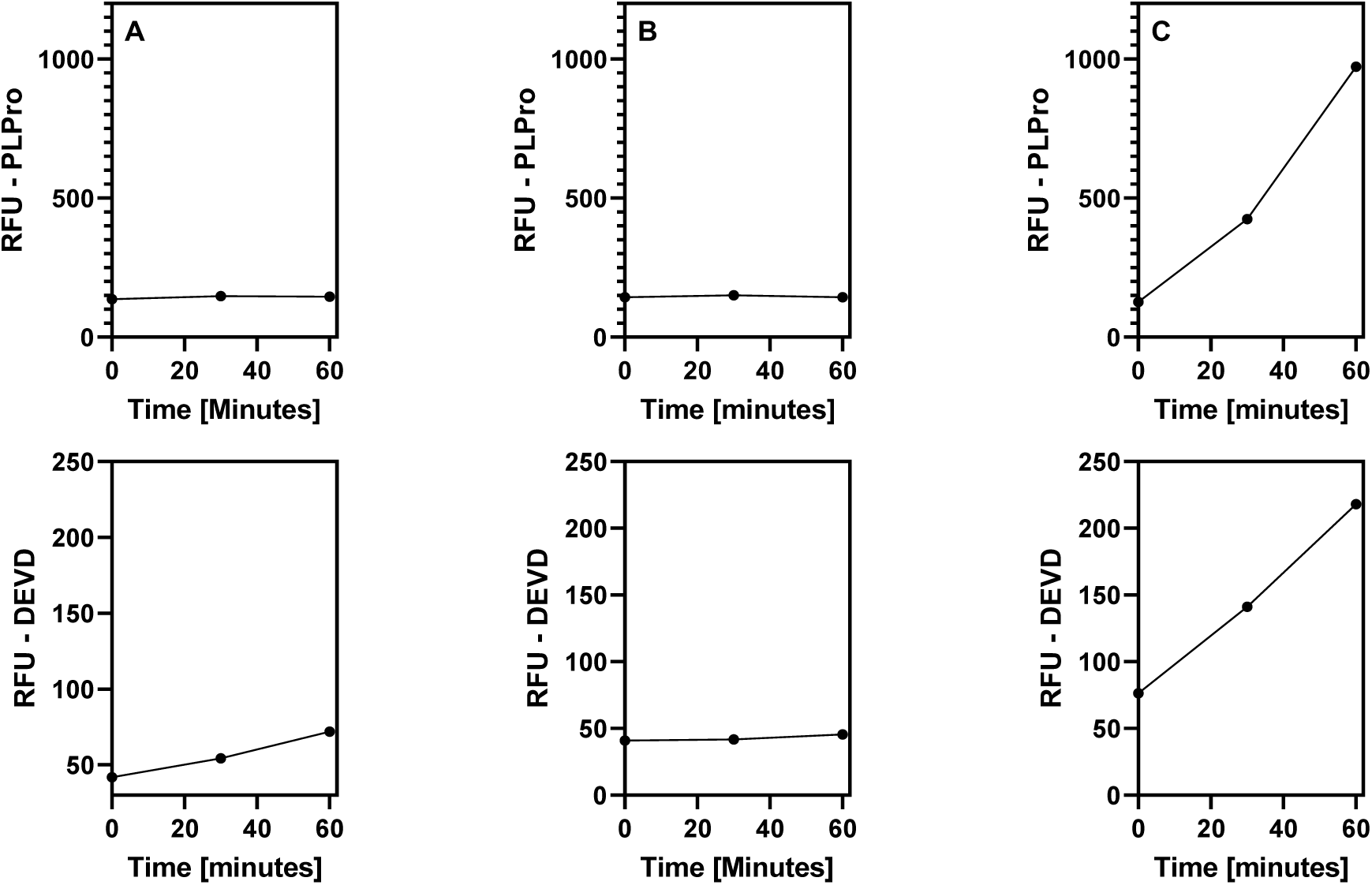
Examples of test results from subject saliva lysates. Specimens were processed as described in Figure 8 and relative fluorescent units (RFU) measured from a vial containing both the PLpro peptide and DEVD peptide (internal processing control). **A)** Negative result: The relative fluorescence units (RFU) from PLpro cleavage activities (top) do not noticeably increase over 60 minutes whereas the RFU from cleavage of the internal processing control peptide do (bottom); the results indicates that the subject was not actively infected with SARS-CoV-2. **B)** Invalid result: RFU from both the PLpro and internal processing control peptides do not increase. The test is invalid. **C)** Positive result: The RFU from both PLpro and internal processing control peptides increases over 60 minutes. This result indicates that the subject was infected with SARS-CoV-2.

The RFU from the Quanti Vial revealed that lysates from saliva contained approximately twice as much material based on nucleic acid content compared to lysate from tongue scrape (4,347 RFU versus 2,545 RFU, respectively). In 6 cases, tongue scrape samples and in 4 cases saliva samples did not contain sufficient material according to the internal processing control, DEVD, using as cutoff a ratio of <10 (delta RFU PLpro/RFU QuantiVial)*100. The first 13 tongue scrape samples that were collected by the same operator included 5 with insufficient material whereas the last 11 samples included only 1 with insufficient material (Table 1 in Supplemental Information), indicating that operator training (more rigorous, yet still painless scraping) should be improved.

**Table 1.** Summary of results obtained from parallel testing of saliva samples with the SARS-CoV-2 protease test and LIAT RT-PCR in patients suspected of COVID-19.

|  |  | LIAT RT-PCR |  |  |
| --- | --- | --- | --- | --- |
|  |  | Positive | Negative | Total |
| PLpro<br>Ratio delta<br>RFU | Positive | 7 | 6 | 13 |
|  | Negative | 2 | 4 | 6 |
|  | Total | 9 | 10 | 19 |
| Positive Agreement |  | =78% (7/9) |  |  |
| Negative Agreement |  | =40% (4/10) |  |  |

### Agreement between the standard of care RT-PCR and protease test

In this first study, the negative percent agreement (NPA) and positive percent agreement (PPA) between the protease test and clinical LIAT RT-PCR were low: The PPA was 45.5% and 86% for saliva and tongue scrape, and the NPA 50% and 33%, respectively. This result was not unexpected given that different specimen types were used for RT-PCR (nasal mid-turbinate) and the protease test (saliva and tongue scrape) that may have had different viral loads.^39^ Further, there were delays in collecting specimens for the protease test after RT-PCR results were returned: 6 samples were collected 0-10 hours after RT-PCR results were returned, 14 were collected between 10-24 hours later, and 3 were collected after more than 24 hours (Supplemental Information, Tables 1 and 2). It is plausible that infections may have cleared in the RT-PCR positive cohort and were not detectable by the protease test, or infections had not yet been detected in the RT-PCR negative cohort but were detectable with the protease test. To better control for these variables—varying specimen types and collection times—a second clinical study was initiated using only saliva specimens that were split for testing with the SARS-CoV-2 protease assay and in-house RT-PCR.

**Table 2.** Summary of results obtained from parallel testing of saliva samples with the SARS-CoV-2 protease test and endpoint RT-PCR in patients suspected of COVID-19. The agreement was significant (p= 0.0073) using Chi square with Yates correction and 0.0103 using Fisher’s exact test (2-tailed or 1-tailed).

|  |  | Endpoint RT-PCR |  |  |
| --- | --- | --- | --- | --- |
|  |  | Positive | Negative | Total |
| PLpro<br>Ratio delta<br>RFU | Positive | 14 | 0 | 14 |
|  | Negative | 2 | 3 | 5 |
|  | Total | 16 | 3 | 19 |
| Positive Agreement |  | =87.5% (14/16) |  |  |
| Negative Agreement |  | =100% (3/3) |  |  |

### SARS-CoV-2 PLpro and MPro activities relative to RT-PCR in saliva

In a second study, 22 patients with COVID-19 like symptoms were prescreened with LIAT RT-PCR performed on nasal mid turbinate specimens at the University of New Mexico Emergency Care Unit. Saliva (500µl) was collected on the same day the LIAT RT-PCR result was returned from 10 patients who tested positive and from 10 patients who tested negative. Two enrolled patients were not able to produce sufficient saliva and were excluded from the study. Each specimen was divided, and one half used for protease testing. RNA was extracted from the other half and analyzed using real time loop-mediated isothermal amplification (RT-LAMP) and endpoint RT-PCR with subsequent gel electrophoresis. Two patients with discordant results were sampled again 5 days later.

The protease tests were performed in 384-well plates and fluorescence was measured in a florescence plate reader. The delta RFU, and the ratios (delta RFU_protease_ /RFU _Quanti dye_)*100 were calculated using GraphPad Prism. The results are shown in Table 3. One sample did not contain sufficient material according to the LoD determined by Quanti dye (patient 19b, see Table 3) and one patient (# 13) had consumed a drink containing a blue dye 1 hour prior to providing their saliva sample that may have interfered with the fluorescence measurements. Results from these patients were excluded from further analysis.

**Table 3.** Results of saliva specimen testing. . The results of testing with LIAT RT-PCR, RT-LAMP and endpoint RT-PCR (EP) are shown in comparison to the results of the Mesa Photonics MPro and PLpro tests. The delta RFU from 3 separate experiments with x3 replicates were averaged (Δmean) and the means normalized relative to the mean RFU from Quanti Dye. For MPro, the cutoff values for Δmean, Ratio, and Quanti Dye RFU were 31,787 RFU, 3.4, and 196,858 RFU, respectively and for PLpro 77,403 RFU, 31.1, and 169,888 RFU, respectively. *Fluorescence interference, **Not enough sample.

| Patient | RT-PCR |  |  | Quanti Dye |  | MPro |  |  |  | PLPro |  |  |  |
| --- | --- | --- | --- | --- | --- | --- | --- | --- | --- | --- | --- | --- | --- |
| | LIAT | LAMP | EP | mean | CV | Δmean | CV | Ratio | CV | Δmean | CV | Ratio | CV |
| 1 | NDE | 48 | 32 | 987,217 | 13.0 | 72,128 | 13.0 | 8.1 | 18.37 | 865,061 | 12.0 | 87.6 | 16.4 |
| 3 | NDE | 44 | 34 | 739,414 | 13.1 | 56,135 | 9.0 | 7.6 | 15.90 | 351,941 | 27.0 | 47.6 | 30.0 |
| 4 | 19 | 21 | 190 | 1,921,140 | 14.5 | 269,173 | 53.0 | 14.0 | 54.95 | 1,862,801 | 23.0 | 97.0 | 27.2 |
| 6 | NDE | 44 | 0 | 462,743 | 2.0 | 20,169 | 28.0 | 4.4 | 28.07 | 74,694 | 8.4 | 16.1 | 8.6 |
| 7 | NDE | 34 | 21 | 1,184,149 | 15.3 | 1,389 | 44.0 | 0.1 | 46.58 | 616,779 | 3.0 | 52.1 | 15.6 |
| 8 | 27 | 20 | 104 | 1,523,219 | 2.8 | 27,261 | 2.1 | 1.8 | 3.53 | 1,343,877 | 3.5 | 88.2 | 4.5 |
| 9 | 14 | 18 | 137 | 530,869 | 9.2 | 8,317 | 16.0 | 1.6 | 18.48 | 81,256 | 21.6 | 15.3 | 23.4 |
| 10 | NDE | 42 | 11 | 1,056,333 | 8.9 | 15,731 | 15.0 | 1.5 | 28.67 | 121,272 | 15.0 | 11.5 | 20.5 |
| 11 | NDE | 42 | 3 | 780,281 | 6.6 | 6,717 | 16.5 | 0.9 | 17.76 | 12,037 | 91.3 | 1.5 | 91.6 |
| 12 | 18 | 22 | 54 | 1,504,312 | 2.6 | 42,509 | 1.2 | 2.8 | 2.88 | 1,694,917 | 3.7 | 112.7 | 4.5 |
| 13* | 13 | 16 | 209 | 817,452 | 9.4 | 6,990 | 31.0 | 0.9 | 32.38 | 130,816 | 12.0 | 16.0 | 15.2 |
| 14a | NDE | NDE | 0 | 1,300,441 | 3.0 | 44,856 | 11.0 | 3.4 | 11.41 | 823,884 | 5.0 | 63.4 | 5.9 |
| 14b | ND | 46 | 17 | 1,583,744 | 4.3 | 189,507 | 5.0 | 12.0 | 6.61 | 1,939,194 | 2.7 | 122.4 | 5.1 |
| 15 | 14 | 18 | 68 | 2,493,530 | 4.4 | 183,788 | 8.0 | 7.4 | 9.11 | 3,768,399 | 5.0 | 151.1 | 6.6 |
| 16 | 15 | 18 | 95 | 1,380,316 | 8.1 | 19,103 | 6.3 | 1.4 | 10.32 | 534,755 | 5.3 | 38.7 | 9.7 |
| 17 | NDE | NDE | 27 | 2,230,545 | 3.8 | 26,651 | 11.0 | 1.2 | 11.64 | 752,027 | 4.8 | 33.7 | 6.1 |
| 18 | 16 | 22 | 145 | 2,370,319 | 1.5 | 203,135 | 8.5 | 8.6 | 8.63 | 2,749,004 | 10.0 | 116.0 | 10.1 |
| 19 | 35 | 22 | 92 | 1,014,791 | 8.5 | 17,596 | 31.3 | 1.7 | 32.39 | 87,800 | 26.8 | 8.7 | 28.1 |
| 19b** | ND | 26 | 82 | 137,574 | 10.6 | 4,262 | 9.0 | 3.1 | 13.90 | -116,505 | -33.3 | -84.7 | 35.0 |
| 20 | NDE | 32 | 32 | 1,921,990 | 2.7 | 275,902 | 7.7 | 14.4 | 8.12 | 1,091,985 | 3.7 | 56.8 | 4.5 |
| 21a | NDE | 50 | 11 | 2,059,971 | 2.2 | 321,324 | 7.0 | 15.6 | 7.33 | 3,692,954 | 1.7 | 179.3 | 2.7 |
| 21b | ND | 52 | 20 | 2,296,219 | 2.1 | 364,133 | 3.5 | 15.9 | 4.07 | 3,740,001 | 6.0 | 162.9 | 6.3 |
| 22 | 26 | 22 | 83 | 2,066,863 | 2.0 | 62,769 | 5.7 | 3.0 | 6.00 | 761,528 | 2.3 | 36.8 | 3.1 |

The limits of detection (mean RFU or ratio+2 x stdev) for MPro and PLpro were calculated from 4 representative experiments using samples that were negative with endpoint RT-PCR. The LoD for sample quantity was determined using Quanti dye RFU as described in methods and was ≤ 196,858 RFU and 169,888 RFU for MPro and PLpro, respectively (Figure 1, Supplemental Information). Samples in which the RFU from Quanti dye was below this cutoff were evaluated as invalid.

### Correlation between MPro and PLpro activities in saliva

In this study, the SARS-CoV-2 MPro activities were measured in addition to the PLpro activities, in order to increase the confidence in our data. The correlation between the MPro and PLpro response was significant across 20 samples tested: the Pearson r value=0.8224 [95% Confidence interval 0.6059 to 0.9255], R^2^=0.6764, and P (one tailed) = < 0.0001, indicating a high likelihood that the fluorescent signal increases were a result of peptide cleavage by SARS-CoV-2 proteases rather than peptide cleavage by other proteases that may be active in the lysates (Figure 10).

**Figure 10.**
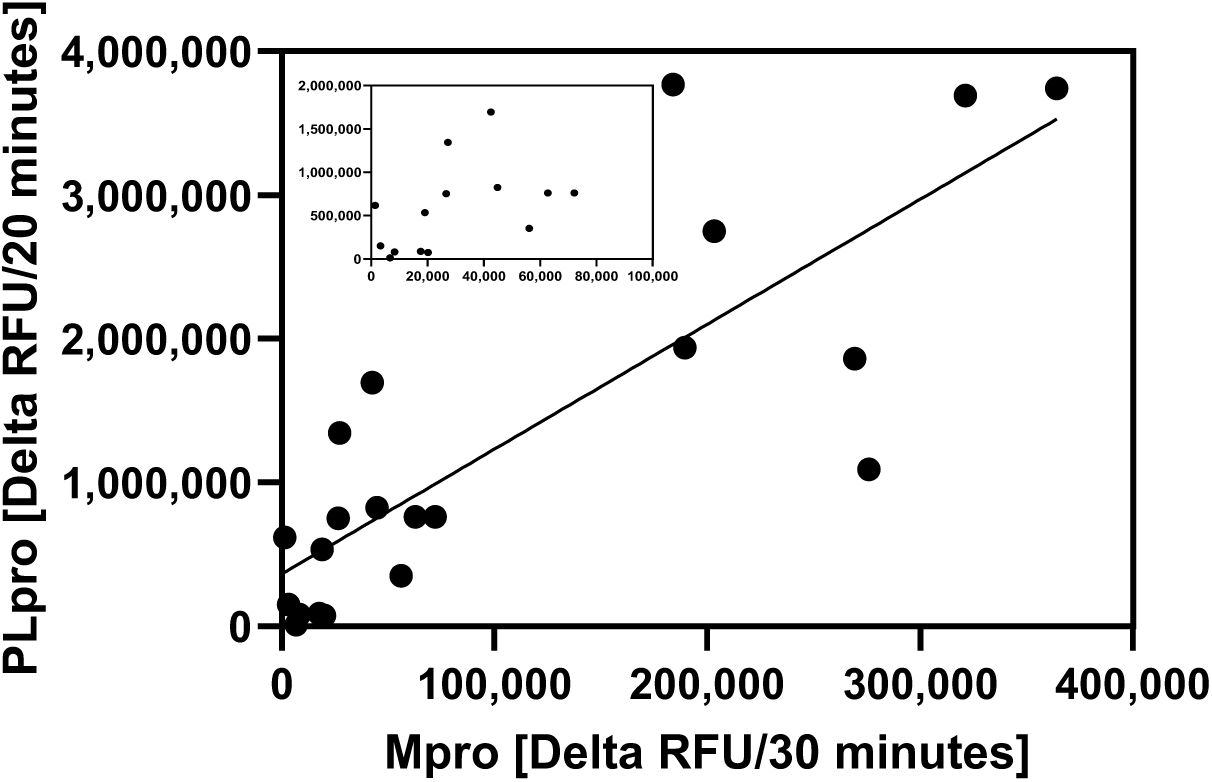
Correlation between SARS-CoV-2 MPro and PLpro activities. The Pearson r correlation between the delta RFU in x2 replicates from the Mpro and PLpro assays from 20 patient specimens was determined using GraphPad Prism. Data points with low RFU values are resolved in the inset.

### Correlation between LIAT RT-PCR, RT-LAMP, and endpoint RT-PCR on NMT and saliva samples

The RT-PCR results using LIAT, LAMP and endpoint RT-PCR agreed for all samples that tested positive with LIAT RT-PCR using a cutoff of < 30 minutes for LAMP and > 50 ratio SARS-CoV-2 amplicon/RNAse P amplicon for endpoint RT-PCR (Table 3). However, in 8 cases that were negative with LIAT RT-PCR, late signals between 32-52 minutes were observed with RT-LAMP. Since with RT-LAMP it cannot be determined whether these late RT-PCR signals resulted from SARS-CoV-2 amplicons or from primer dimers with which the fluorescent dye intercalates, RNA samples were amplified using endpoint RT-PCR and the PCR products separated on an agarose gel. An example of 9 samples (patient IDs 1-11) is shown in Figure 11.

**Figure 11.**
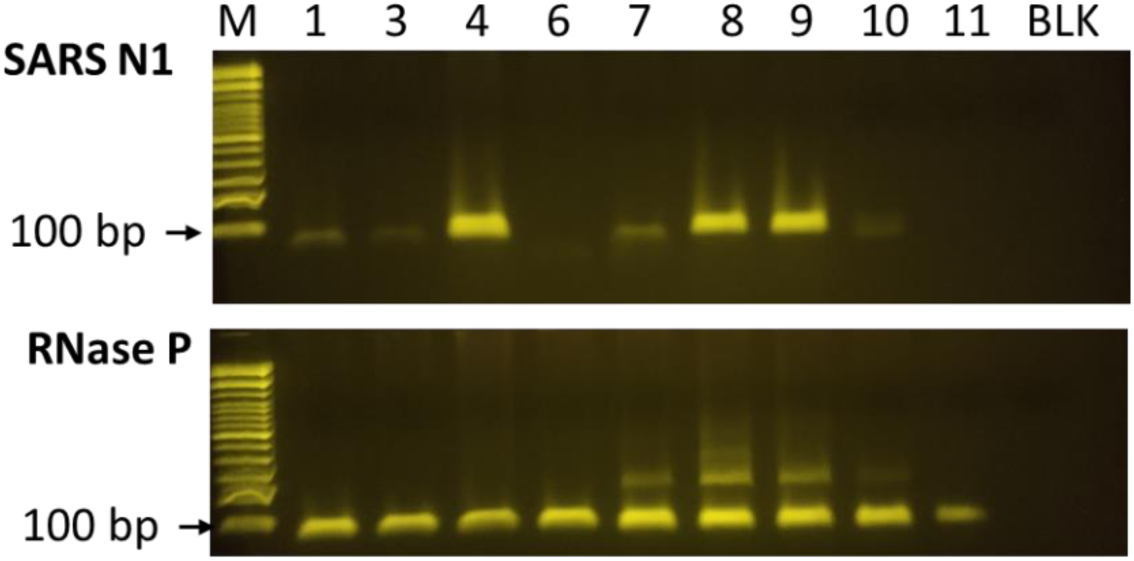
Agarose gel electrophoresis of RT-PCR amplicons. RNA was isolated from saliva specimens and reverse transcribed and amplified using endpoint RT-PCR mix containing SARS N1 primers (top) or RNase P control primers (bottom) and separated on a 2% agarose gel. BLK=reaction without RNA. M=100 base pair ladder, the 100 bp band is indicated by an arrow.

The results show strong amplification of the SARS-CoV-2 N1 gene in 3 samples that tested positive with LIAT and RT-LAMP (patient IDs 4, 8, and 9) and weak amplification in 4 samples that tested negative with LIAT RT-PCR and were late positive with RT-LAMP (patient IDs 1, 3, 7, and 10). 2 samples that were negative with LIAT and RT-LAMP did not contain amplicons (patient IDs 6 and 11). All samples were positive for the human RNAseP gene. To normalize for the amount of amplifiable RNA in the samples, the ratio was calculated between the integrated intensities of the SARS-CoV-2 amplicons and the RNAse P gene (see Table 3).

### PPA and NPA depends on the type of RT-PCR test used as comparator

The negative percent agreement (NPA) and positive percent agreement (PPA) of the SARS-CoV-2 test was calculated relative to LIAT RT-PCR using the normalized delta PLpro RFU (ratio RFU) from Table 3.

The sensitivity of detection was high, but the specificity was low due to 6 positives that were not detected using LIAT RT-PCR.

In contrast, when endpoint RT-PCR was used as a comparator, the PPA and PPA were 87.5% and 100% (Table 2). One of the two false negative results is from a patient (patient 19) with a high Ct value of 35 in LIAT RT-PCR, suggesting that residual, non-infectious RNA may have been detected.

### Earlier detection of infection with the SARS-CoV-2 protease test than with LIAT RT-PCR

One sample (14a) that tested negative with all three RT-PCR tests exhibited very strong delta RFU from the MPro and PLpro peptides (Table 3). The subject provided another sample 5 days later and the specimen (14b) was retested, and showed a positive result in endpoint RT-PCR. The normalized PLpro and MPro protease activities in sample 14b were approximately 2-fold higher than those obtained previously in sample 14a (3.4 and 12 for MPro and 63.4 and 122.4 for PLpro in samples 14a and 14b respectively). Endpoint RT-PCR was low positive in 14b (ratio N1/RNAase P=17). The results indicate that the SARS-CoV-2 protease test may have detected an infection before extracellular viral fragments were produced in quantities that could be measured by RT-PCR. Likewise, serial testing of patient 21, who provided a second sample after 5 days showed a high level of SARS-CoV-2 protease activity on both days of testing, in absence of a positive LIAT RT-PCR result, but with late signals in RT-LAMP (50 and 52) and endpoint RT-PCR (ratio N1/RNAase P=11 and 20).

The normalized ratios of PLpro activities can be divided into three groups: 8 samples with high PLpro activity (ratios between 85-179 RFU), 7 samples with moderate PLpro activity (ratios between 24-63 RFU) and 5 samples with low PLpro activities (ratios between 2-17 RFU), of which 3 samples tested negative with endpoint RT-PCR. The two presumptive false negatives are included in the positive cohort. The distribution is shown in a box and whisker plot (Figure 12).

**Figure 12.**
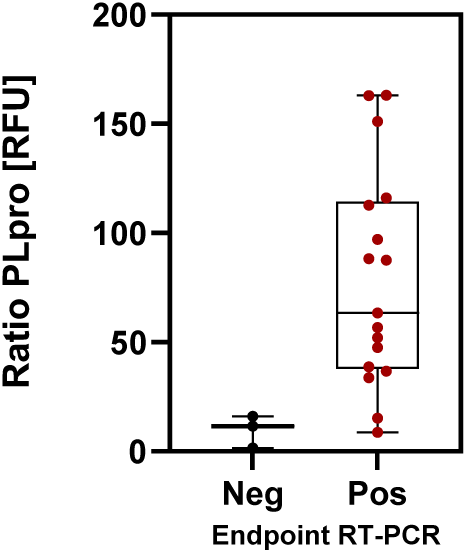
**Box** and whiskers plot of mean, max and min of PLpro ratios in a negative (Neg) and positive (Pos) cohort with endpoint RT-PCR as a comparator.

## DISCUSSION

In this work, we have shown feasibility of a SARS-CoV-2 PLpro activity-based assay as a rapid and simple diagnostic test for detecting active infections in patient saliva, potentially earlier and more accurately than is possible with diagnostic RT-PCR. The test was performed in a POCT site by minimally trained operators or automated in a laboratory setting using a fluorescence plate reader. The specificity of SARS-CoV-2 PLpro peptide cleavage was supported by data from VeroE6 cells infected with SARS-CoV-2. In addition, we demonstrate the application of the platform for screening of antiviral targeted drugs and neutralizing agents at lower time and cost and greater ease of use compared to current viral proliferation assays.

### PLpro activity as a marker of SARS-CoV-2 infection

To our knowledge, using PLpro activity as a marker of infection is a novel approach for SARS-CoV-2 diagnostics. A diagnostic test by Todos Medical using SARS-CoV-2 MPro was announced but appears to not have been further developed.^40^ Detecting PLpro activity in crude, unprocessed cellular lysates was successful due to a rigorous design and optimization of the test platform to ensure that the reporter peptide was cleaved predominantly by SARS-CoV-2 PLpro and not significantly by papain-like proteases from other human coronaviruses or by other proteases that are present in crude lysates. The specificity of cleavage was accomplished by i) using fluorophores that fluoresce only when the bond between the terminal amino acid and the fluor is cleaved, whereas any potential cleavage by other proteases between upstream amino acids does not result in significant fluorescent signals; ii) increasing the sensitivity of detection by designing 16-mer and 74-mer peptides composed of amino acids from the ISG15 protein that PLpro is known to preferentially bind and cleave.^31^ In this study ISG15-based peptides resulted in a 43-fold increase in sensitivity in biochemical assays compared to the minimal tetrapeptide. In addition, the background cleavage of ISG15-based peptides was low, likely because only few deISGylating proteases are expressed in human cells that would cleave the ISG15-based peptides of the test, e.g., the ubiquitin-specific proteases USP5, USP16, and USP18.^41,42^ Accordingly, we observed that cleavage of the PLpro ISG15-based peptide Pun74 by host cell proteases in lysates from uninfected VeroE6 cells was less than 10% relative to the cleavage in lysates from infected cells; iii) increasing the specificity of detection by incorporating unnatural amino acids into the LXGG cleavage site of the peptide. Specific cleavage of the ISG15-based peptides by recombinant PLpros from SARS-CoV and SARS-CoV-2 was observed, whereas no significant cleavage was observed by recombinant papain-like proteases from the other human coronaviruses (HCoVs MERS, OC43, NL62, 229E, and HKU1) or by the deISGylating human USP18 protease.^32^

### Comparison of saliva and tongue scrape specimens in a PoC test

The test was performed by minimally trained operators using a battery-operated fluorometer and without a need for precision volumetric control, thus making the test compatible with use in remote and hard to reach locations. Fluorescence increases were measured for 60 minutes, although 30 minutes was shown to be sufficient. In this POCT study, dual fluorescence from an internal processing control peptide (DEVD_2_-R110) and the SARS-CoV-2 PLpro peptide (Pun16-AMC) was monitored in the same sample vial. Unfortunately, 26% (6 out of 23 samples) of the tongue scrape specimens did not contain sufficient amount of material according to the cutoff value set for the internal processing control DEVD. The high failure rate of tongue scrapes could be explained by an initial hesitancy of the operator to scrape vigorously as the majority of invalid samples were obtained during the first half of the study and the specimen quantity increased with operator experience. Saliva had a lower rate of invalid specimens (17%, 4 out of 23 samples), but some patients had difficulty producing the amounts of saliva required for the test (0.5 ml), especially older patients and those taking certain vitamin supplements and medication. In those cases, and for young children, tongue scrape specimens deserve future consideration.

Several limitations of this study must be noted. Our sample size was limited, and the pretest probability was high since only cases with strong clinical signs of COVID-19 were included. More than half of the specimens for the protease test were collected 10 to 48 hours after results from RT-PCR were returned, and unsurprisingly, the agreement with RT-PCR performed on nasopharyngeal swabs was low (PPA=86% and 46%, NPA=33% and 50% in tongue scrapes and saliva, respectively). It is plausible that infections in the RT-PCR positive cohort had cleared before specimens were collected for the protease test or had not yet been detected in the RT-PCR negative cohort in which 5 patients tested positive with the protease test. Further, difference specimen types were used—nasopharyngeal swabs (NPS) for RT-PCR and saliva for the protease test—that reportedly carry different viral loads.^39^

### SARS-CoV-2 RT-PCR in saliva

To minimize inconsistencies between RT-PCR and PLpro results due to the specimen type and collection time, we conducted a second pilot study, in which saliva specimens were collected on the same day that diagnostic RT-PCR results were returned. The saliva was split for SARS-CoV-2 protease testing and for in-house RT-PCR. We chose real time RT-LAMP since its performance in saliva was reported to equal that of RT-PCR in a workplace surveillance study.^43^ However, a drawback of real time RT-LAMP is that late signals resulting from true SARS-CoV-2 amplification products cannot be distinguished from primer dimers that readily form due to the large number of primers used in LAMP (6 per target) and their length (40-50 bases).^44^ To distinguish between those artefacts and SARS-CoV-2 amplicons, endpoint RT-PCR was performed using SARS-CoV-2 N1 and internal control RNAse P primers with sequences corresponding to those recommended in 2020 by the CDC for SARS-CoV-2 testing. The products were separated by gel electrophoresis and the ratio of SARS-CoV-2 amplicons relative to control amplicons calculated. This may have been an imperfect analysis since the positive control primers originally recommended by the CDC are located in the same exon and cannot distinguish whether the amplicons were derived from RNA or DNA, i.e., false negatives may have been obtained from samples in which RNA had degraded.^28,45^ The agreement between the three RT-PCR test was 100% when a Ct value of ≤ 30 minutes was selected for RT-LAMP and a ratio N1/RNAse P ≥50 for endpoint RT-PCR. However, in 8 cases, late signals ≥ 30 minutes were observed with RT-LAMP and ratios between N1/RNAse P ≥ 20 for endpoint RT-PCR in 5 samples that were negative with LIAT RT-PCR.

The dramatic differences of positivity between LIAT RT-PCR (10 positives of 20) and endpoint RT-PCR (16 positives of 20) deserves further consideration. One explanation for the divergent results is that with LIAT RT-PCR samples with cycle threshold (Ct) values above 35 are reported as negative whereas we performed endpoint RT-PCR for 40 cycles, which may have increased the numbers of positives. Another factor is that endpoint RT-PCR was performed on RNA isolated from saliva, whereas LIAT was performed using nasal swabs. Reports show that there is a high degree of variability in detecting SARS-CoV-2 RNA among different specimen types that were collected at the same time from the same person.^46^ One report showed a RT-PCR test positivity rate of 52.6% in nasopharyngeal swab specimens compared to 92.14% in saliva from the same patients, as well as higher viral loads and prolonged shedding.^39^

### Positive and Negative Agreement between SARS-CoV-2 ABDx assays and RT-PCR

When the results of this study were compared to those from endpoint RT-PCR, the sensitivity was 93.8 % (15 of 16) and the specificity 100%, whereas the sensitivity was 88.9% and the specificity 40% when using diagnostic LIAT RT-PCR as a comparator (Tables 1 and 2). Several arguments can be made in support of the protease test results: 1) the normalized ratios >24 of MPro and PLpro activities corresponded with the results from endpoint RT-PCR (15 positives out of 19 specimens included in the study); 2) A significant correlation was measured between the SARS-CoV-2 PLpro and MPro proteases (Pearson r=0.8224, Figure 10); 3) Although the study included only a small number of patients, the high number of positives detected is plausible because the study took place during the Omicron epidemic wave in New Mexico from the beginning of June to the end of September 2023 when test positivity trends peaked at 14.3%.^47^ Further, the study included only patients with severe COVID-19 symptoms requiring emergency medicine services, thus increasing the pre-test probability of COVID-19.

Only one presumptive false negative result was observed in the protease test relative to endpoint RT-PCR (patient 19 with a PLpro ratio=9 versus ratio endpoint RT-PCR=159). This patient had a late CT value of 35 with diagnostic LIAT PCR and may no longer have been harboring an active infection, as indicated by the low SARS-CoV-2 PLpro activity. One presumptive false positive result relative to endpoint RT-PCR was observed in patient 14 (normalized ratio PLpro=63.4 versus endpoint RT-PCR ratio =0; see Table 3) but serial testing after 5 days strongly indicated that the PLpro test had accurately detected the presence of an infection; the normalized PLpro ratio doubled from 63.4 to 122.4 and an amplicon was detected in endpoint RT-PCR in the second specimen collected (ratio N1/RNAseP=17).

### Limitations

This study included only a limited number of clinical specimens with a high COVID-19 pre-test probability as a proof-of-concept for the performance of the SARS-CoV-2 activity assay. A substantially larger set of diverse clinical samples is needed to reliably estimate the sensitivity and specificity of the assay. Additionally, the small set of serial samples from patients with incongruent results is only an indication that the PLpro ABDx assay might detect SARS-CoV-2 infections earlier and within a narrower window than RT-PCR. If verified, earlier and more accurate detection would have tremendous implications for patient management and infection control.

### SARS-CoV-2 PLpro activity platform for detection of any variant

It is expected that the PLpro activity assay will detect any SARS-CoV-2 variant. Although mutations in PLpro do occur, they do not affect the catalytically active regions which would produce a false negative protease test result because this would interfere with viral survival. Mutations are reported in other areas of PLpro that may affect substrate binding.^19^ However, our reporter peptides are 17 and 74 amino acids in length and unlikely to lose their ability to bind to the enzyme substrate binding site in the event of single amino acid substitutions. Future testing of variants in cultured cells is planned to examine this.

### Application of SARS-CoV-2 ABDx platform for antiviral screening

The data presented here also support the application of the protease platform as a screening tool for antiviral drugs. In our study, the inhibition value for the PLpro inhibitor GRL0617 in VeroE6 cells was 38.3µM (Figure 6), which is in close agreement to the effective inhibitory concentration of 21 ± 2 μM reported in a cell-based assay using cytopathic effect (CPE) as readout.^34^ Currently, only 2 oral antiviral drugs are approved for use in the United States: Nirmatrelvir (a component in Paxlovid) that targets MPro, and molnupiravir, a nucleoside analog developed by Merck targeting RNA-dependent RNA polymerase (RdRp).^48,49^ Over the past 3 years variants with mutations in MPro have caused drug resistance under therapeutic pressure while concerns have been raised about molnupiravir’s capacity to trigger SARS-CoV-2 mutations.^50–52^ It is therefore important to study additional therapeutic targets, such as SARS-CoV-2 PLpro. Inhibiting the dual functions of PLpro—suppression of the innate immune response by cleaving ISG15 and processing of the viral polyprotein—have prompted drug development efforts, but to date, none of the promising drug candidates have advanced to animal models.^25,53,54^ One explanation is that most biochemical studies examined only the minimal proteolytic domain of PLpro without considering the other domains in SARS-CoV-2 proteins that influence PLpro activity and function.^25^ Although this limitation is overcome by tests that measure the extent of drug-induced cytopathic effect (CPE) in cells or quantify the amount of viral RNA in response to inhibition, these phenotypic assays take days to complete and require several user steps. Assays using genetically modified SARS-CoV-2 strains or replicons are faster and simpler but are suitable only for the virus strains they have been established for.^55^ In contrast, the SARS-CoV-2 PLpro assay described here is a simple add-and-measure homogeneous assay that can be scaled up to run in 96-well or 384 well plates. Time course data demonstrates that SARS-CoV-2 PLpro activity or inhibition can be detected in as little as 6 hours after infection with 0.1 MOI (Figure 5) and testing can be completed in one working day with minimal user steps. Including caspase-induced cleavage of the DEVD peptide in the same well as PLpro peptide cleavage can normalize for the amount of cells/lysate present and provide additional information on cell health. In fact, one study relied solely on monitoring caspase activity to determine the activity of anti-SARS-CoV-2 antivirals and neutralization.^55^

The assay platform also shows promise for detecting new neutralizing antibodies (Figure 7) that are urgently needed to combat the loss of efficacy of previously effective monoclonal therapeutics due to SARS-CoV-2 variants, as manifested by increased numbers of breakthrough infections in vaccinated individuals.^56–60^ Our data demonstrates successful detection of neutralizing activity from patient serum and promises to be a rapid, low-cost alternative to other multi-step and laborious phenotypic assays, such as plaque reduction neutralization test (PRNT), fluorescent antibody virus neutralization (FAVN) assays, or immunological assays.^26^

## Conclusion

Functional activity-based diagnostic assays such as the SARS-CoV-2 PLpro assay presented here are gaining increasing importance as reliable tools for detecting pathogen infections. Significant advantages are that test reagents are inexpensive and can be synthesized in bulk for widespread distribution with minimal turnaround times. The cost efficiency and scalability of the test make it suitable to test asymptomatic and presymptomatic carriers in screening efforts. Further, as indicated by our data, the SARS-CoV-2 protease test detects only active infections and possibly earlier than with RT-PCR. Early and accurate determination of whether a person is actively infectious is a critical factor in making treatment and quarantining decisions. The expected resistance of SARS-CoV-2 PLpro to mutations in emerging variants makes it a powerful surveillance tool to prepare for future outbreaks. Further, we demonstrate the potential of the assay platform for rapid and homogeneous cell-based screening of antiviral therapeutics against SARS-CoV-2.

This report encourages the application of the platform as an effective diagnostic for other infectious diseases, such as influenza, viral Hepatitis C, or mycobacterial tuberculosis, through careful selection of an appropriate enzyme and the design of a peptide that it specifically cleaves.

## Data Availability

All data produced in the present study are available upon reasonable request to the authors

## Acknowledgements

We gratefully acknowledge technical assistance provided by Los Alamos National Laboratory through the New Mexico Small Business Assistance (NMSBA) program. We thank Sarah Lavelle, Munia Omer and Kate Browning (University of New Mexico) for support with IRB submission and active recruitment.

## Author Contributions

FHR conceived the project, obtained funding, performed the research, and wrote the original draft. AGJ and LIM collected specimens and performed clinical testing at UNM. KW and YMS performed research. SBB conceptualized and oversaw BSL3 experiments and CY performed experiments with live SARS-CoV-2 at UNM. BB oversaw experiments relating to cell lysis and SARS-CoV-2 inactivation by lysis buffer. NV performed experiments with biosafety level 2 HCoVs. JTB and SCB led the clinical trial administration efforts. WJB supported data analysis and writing the original draft.

