## Supplemental Data 1 for "A Cell-Based Papain-like Protease (PLpro) Activity Assay for Rapid Detection of Active SARS-CoV-2 Infections and Antivirals"

**Supplemental Information**

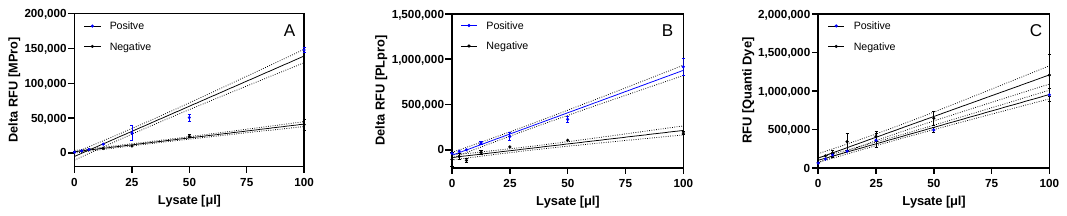

**Figure 1. Limit of Detection (LoD) determined by Quanti Dye.** Serial dilutions of lysates from patients who tested positive or negative for SARS-CoV-2 by RT-PCR were prepared in lysis buffer. A) Peptides cleaved by MPro or B) PLpro were reacted for 30 minutes with lysates. C) Quanti Dye was added to lysate in separate wells. Linear regression of the delta RFU was performed using GraphPad Prism and the y intercepts of the reaction from positive (b2) and negative (b1) lysates calculated using the formula x= b2-b1/m1-m2, where b=intercept and m=slope. The LoDs for the MPro and PLpro substrates were 6.4µl and 3.9µl lysate, respectively. Interpolation of these values to the linear fit from the Quanti Dye RFU was performed using the formula y=mx+b and resulted in LoDs for Quanti Dye of 196,858 RFU for MPro and 169,888 RFU for PLpro peptides, respectively.

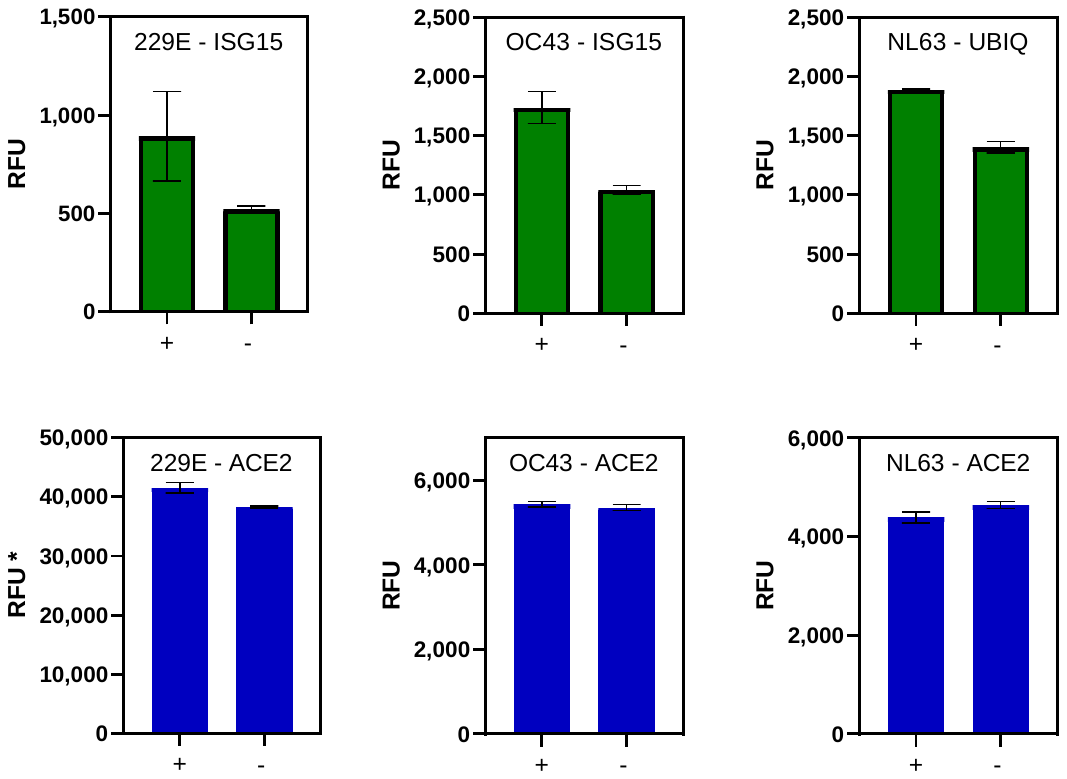

**Figure 2. DUB activities in lysates from cells infected with human coronaviruses. Top.** Cells (~100,000) were infected (+) with 2 MOI (229E and OC43) or 1 MOI (NL63) and incubated for 24 hours. Control are uninfected cells (-). Lysates were prepared and incubated with ISG15-AMC or Ubiquitin-AMC and fluorescence monitored after 15 minutes. **Bottom**. Positive control peptides that are cleaved by ACE2 were added to separate wells containing lysates, demonstrating comparable quantities of active lysates from infected and uninfected cells.

***** the measurement was performed with higher instrument gain settings

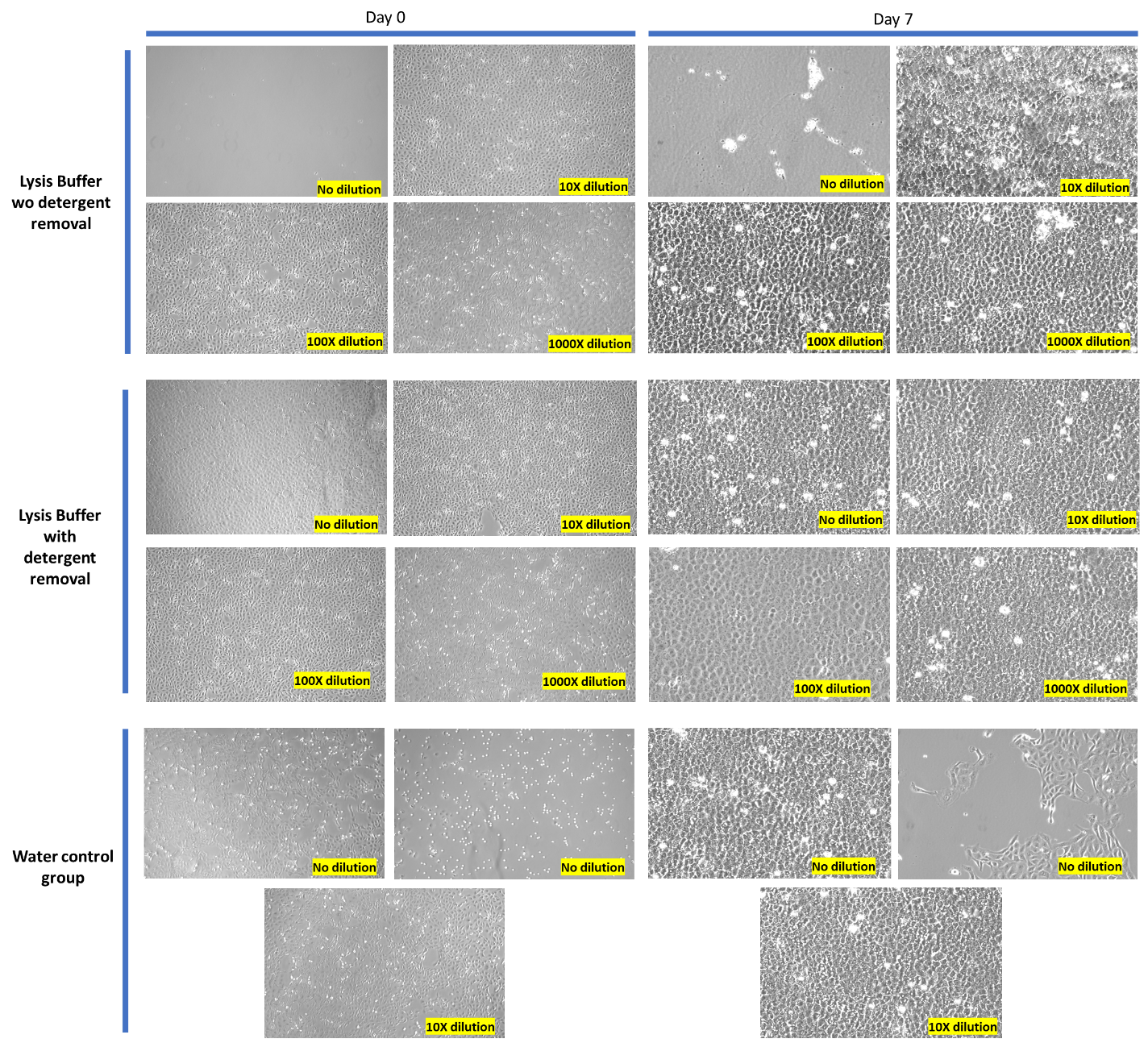

**Figure 3 –** **Host cell lysis by Mesa Photonics lysis buffer.** Representative microscopy images at 200X magnification for a cytotoxicity assessment of lysis buffer. Vero-E6 cells exposed to undiluted lysis buffer experienced significant cell death on the day of treatment. Dilutions of the lysis buffer at 1:10 or greater in viral growth media resulted in no observed cell death on both the day of treatment and 7 days post-treatment. Furthermore, when a detergent removal step was implemented, cells treated with undiluted lysis buffer exhibited no cytotoxic effects during the experiment period. Cells treated with water did not exhibit cytotoxicity, although limited small regions of the well displayed cell detachment which may be due to cell plating. However, cells began to regrow in these areas during 7 days of experiment, suggesting that the diluent used to prepare the lysis buffer did not cause cytotoxic effects.

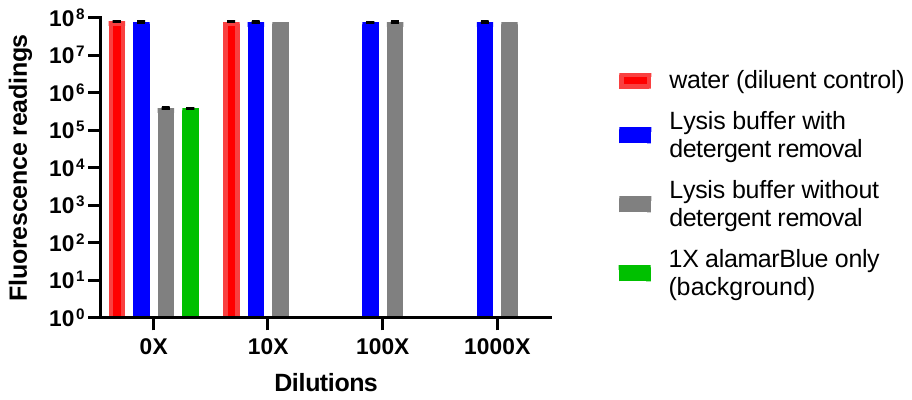

**Figure 4. Host cell lysis by lysis buffer determined by cell viability staining**. Cell viability fluorescence signals (Y-axis) were obtained from test and control groups. The x-axis denotes the dilution in viral growth media of lysis buffer, detergent-removed lysis buffer, and water (diluent control). The gray columns represent cells treated with lysis buffer without filtration through PDRSC; the blue columns correspond to cells treated with lysis buffer filtered through PDRSC; the red columns represent cells treated with water; and the green column represents 1X alamarBlue only group (background control). Two measurements were taken for each dilution in every sample group and 4 measurements for the 1X alamarBlue only control.

Cells exposed to undiluted lysis buffer exhibited a 2-log reduction in absolute fluorescence signal compared to the other groups, and a similar fluorescence reading compared to 1X alamarBlue only background control group. This indicates complete host cell lysis which is consistent with the microscopic examination observations.

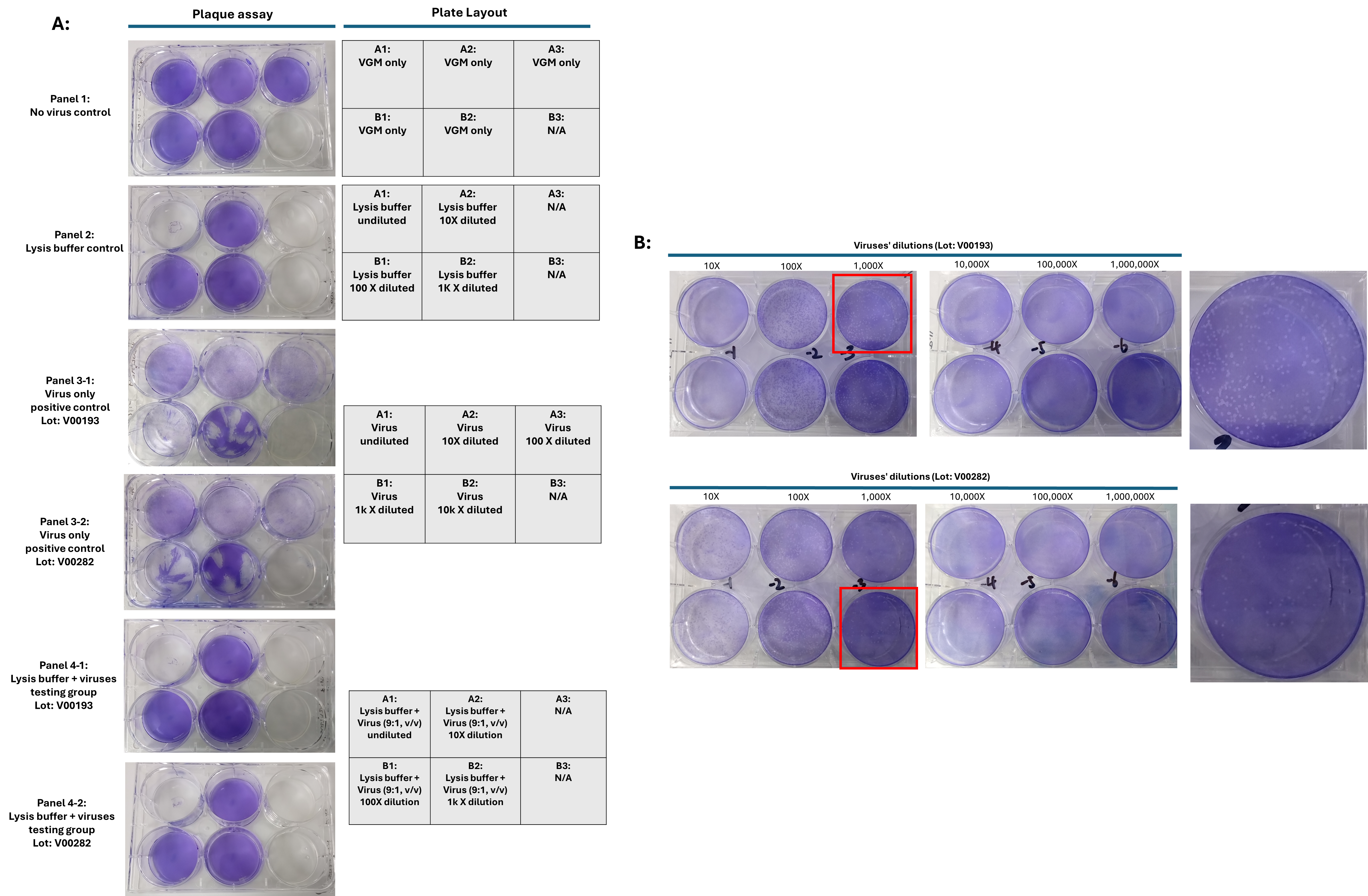

**Figure 5 - Lysis buffer inactivation efficacy against SARS-CoV-2.** Various dilutions of SARS-CoV-2 from two different lots were prepared from viral stock and combined with lysis buffer or VGM in a ratio of 1:9. Retained virus infectivity was determined by plaque assay. **A. Comparison of plaque formation in different treatment groups**. Healthy cell growth was observed in the no-virus control group (**panel 1**), whereas the group treated with undiluted lysis buffer (with or without virus) demonstrated significant cell death (**wells A1 in panel 2, 4-1 and 4-2**), whereas wells treated with diluted lysis buffer maintained a healthy cell monolayer (**wells A2-B3 in panel 2**). The virus-only control group (**panels 3-1 and 3-2**) formed plaques across all virus dilutions (from undiluted to 10,000-fold dilution). In contrast, the group treated with virus in lysis buffer (wells A2-B2 in panels 4-1 and 4-2) showed no plaques, indicating effective inactivation of the virus by lysis buffer. The well with undiluted lysis buffer showed no crystal violet staining (**well A1 in panels 2, 4-1 and 4-2**) due to lysis buffer cytotoxicity. **B. Repeat of virus positive control in plaque formation assay:** To obtain better defined and countable plaques, we repeated the plaque assay using SARS-CoV-2 from the lots we tested in A with a higher concentration of CMC (3%) for sharper plaque formation. As shown in figure B, sharp and countable plaque are formed in infected VeroE6 cells treated with 1,000X-10,000X dilutions of virus. The titer of the virus lot V00193 was calculated as 4.2 X10^6 pfu/mL and the titer of virus lot V00282 was calculated as 9.8 X10^5 PFU/mL. The combined data suggest that lysis buffer is able to fully inactivate SARS-CoV-2, at least within these infectious titers, and that the virus inactivation effect is independent of the virus lot.

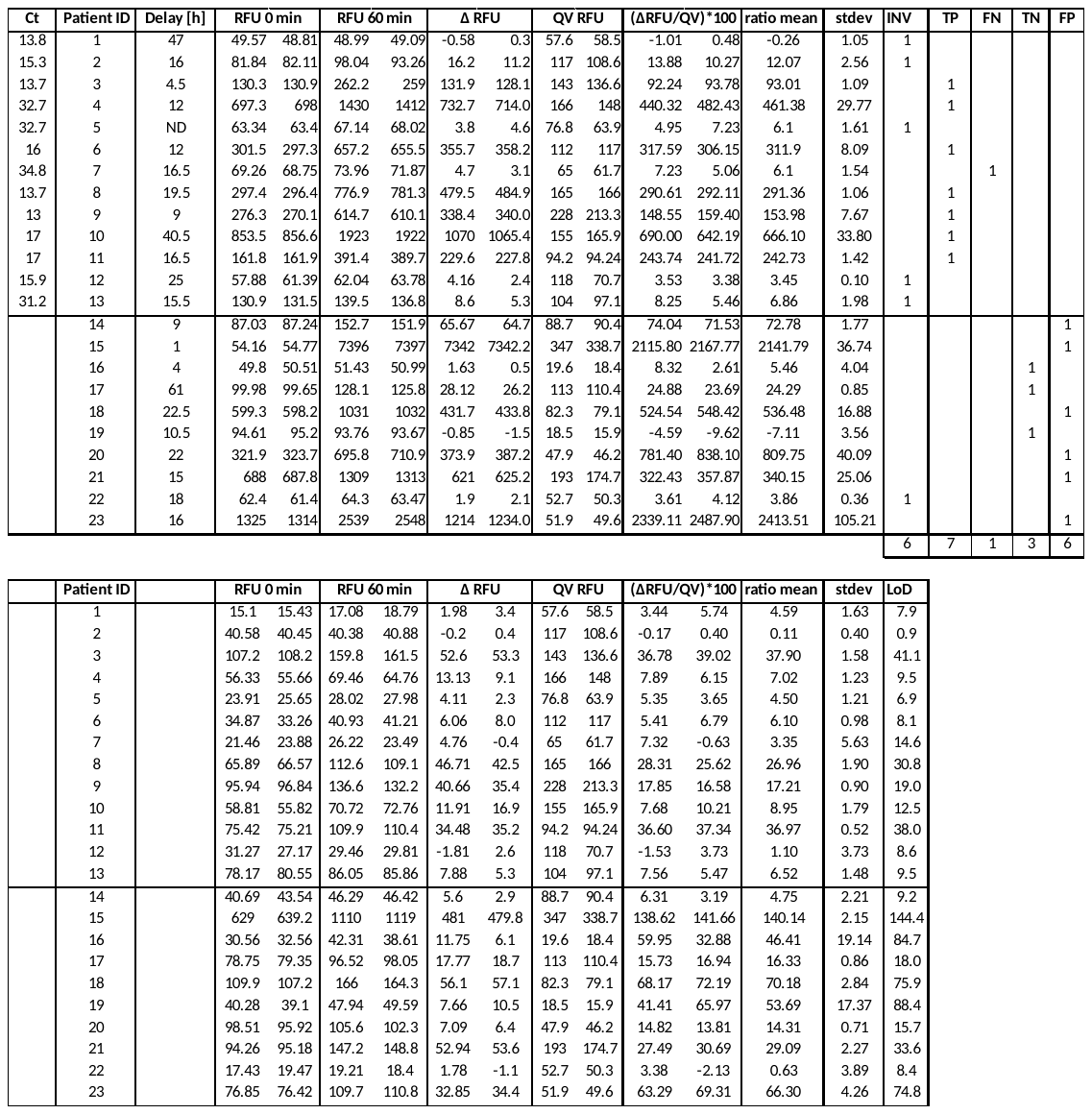

**Table 1. Results from PoC testing for SARS-CoV-2 infection in lysates from tongue scrape specimens from 23 patients**. The cycle threshold (Ct) values obtained for 23 patients and the delay (hours) between return of diagnostic RT-PCR and sample collection are given. **Top**. PLpro. Relative fluorescence units (RFU) between the 60 minute and 0-minute reads PLpro activity and the delta RFU (Δ) are shown. The ratio between delta RFU and Quanti Vial (QV) was calculated and the mean determined. Invalid samples (INV), true positives (TP), false negatives (FN), true negatives (TN), and false positives (FP) are indicated. **Bottom**. DEVD. The ratio of delta RFU was calculated and the LoD detection determined as <10 ratio mean.

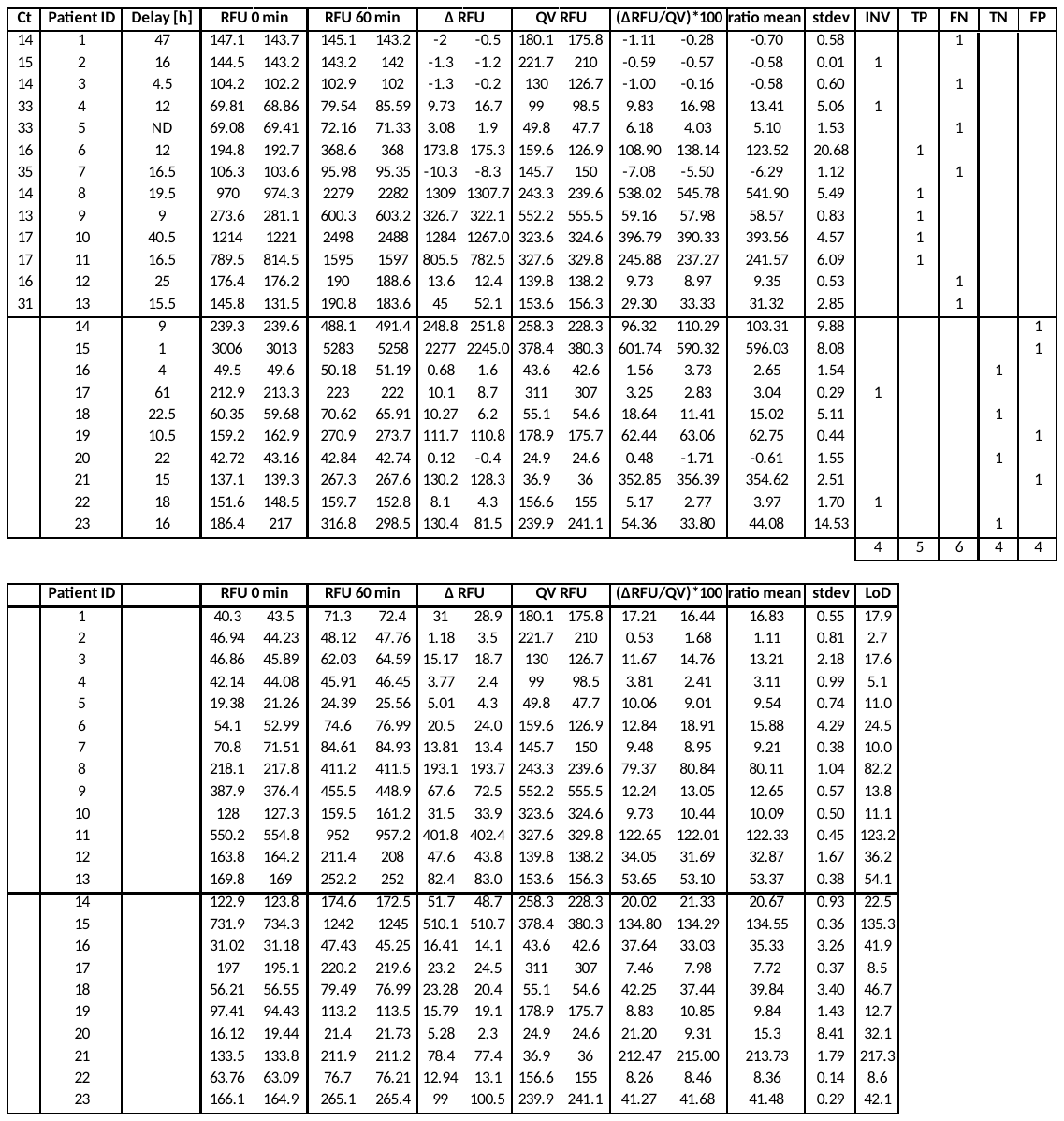

**Table 2. Results from PoC testing for SARS-CoV-2 infection in lysates from saliva specimens from 23 patients**. The cycle threshold (Ct) values obtained for 23 patients and the delay (hours) between return of diagnostic RT-PCR and sample collection are given. **Top**. PLpro. Relative fluorescence units (RFU) between the 60 minute and 0-minute reads of PLpro activity and the delta RFU (Δ) are shown. The ratio between delta RFU and Quanti Vial (QV) was calculated and the mean determined. Invalid samples (INV), true positives (TP), false negatives (FN), true negatives (TN) and false positives (FP) are indicated. **Bottom**. DEVD. The ratio of delta RFU was calculated and the LoD detection determined as <10 ratio mean.
